# Defining Manuscript Peer Review: An International Delphi Consensus Study

**DOI:** 10.64898/2026.10.05.26364760

**Authors:** Mehvish Masood, Ka Hin Tai, Julia Soto Rizzato, Janice Tufte, Michael Mittelman, Christine Von Raesfeld, Jessie McGowan, Balazs Aczel, Olavo B. Amaral, Howard Bauchner, Khaoula Ben Messaoud, George Currie, Annette Flanagin, Diego A. Forero, Ann Blair Kennedy, Madhuri S. Kurdi, Geovani López-Ortiz, Gwinyai Masukume, Durga Prasanna Misra, Arvind Babu Rajendra Santosh, Luisa Schonhaut Berman, Abrar K. Thabit, Sadia Yaseen, Alexandra Bodnaruc, Isabelle Boutron, Jocalyn Clark, Martin Offringa, Ludo Waltman, David Moher

**Author notes:** Corresponding Author Details: Dr. David Moher; Ottawa Hospital Research Institute, Ottawa, Ontario, Canada. Email addresses: MMa, KHT, JSR, JT, MMittelman, CVR JM BA, OBA, HB, KBM, GC, AF, DAF, ABK, MSK, GL, GM, DPM, ABRS, LSB, AKT, SY, AB, IB, JC, MO, LW, DM.

## Abstract

**Objectives:** To conduct a Delphi process to inform an international, consensus definition of manuscript peer review.

**Design:** Three-round Delphi consensus study including two surveys and one virtual consensus meeting. Peer review features identified through a prior scoping review were rated on a 5-point Likert scale in Rounds 1 and 2 and voted on using include, exclude, and abstain options in Round 3. Consensus was defined as _≥_80% agreement.

**Setting:** International experts representing interest holders affected by manuscript peer review across the fields of biomedical, health, and social sciences, economics, and healthcare-related industries.

**Participants:** Round 1 participants were purposively recruited via email from corresponding authors of articles included in the preceding scoping review. Round 2 participants were those who completed Round 1. Round 3 participants were purposely selected to address gaps in representation from earlier rounds, with attention to diversity of perspectives and equity-relevant characteristics informed by PROGRESS-Plus (e.g., geographic location, gender, race/ethnicity, occupation, and socioeconomic context). Participants were required to communicate in English.

**Main outcome measures:** Consensus on the terminology, scope, characteristics, functions, processes, and reviewer characteristics that should inform a definition of manuscript peer review.

**Results:** Across the three rounds (N=143, 109, and 16 participants, respectively), 44 peer review features across six domains (*Characteristics, Function, Motivation, Assessment, Process, and Challenges*) and six peer reviewer characteristics met the consensus threshold for inclusion. Consensus was also reached on terminology to describe manuscript peer review and the peer review workflow. Results were synthesized into a final conceptualization of manuscript peer review, consisting of one definition and eight pillars.The consensus definition is: “Manuscript peer review is a structured process by which human reviewers assess scholarly manuscripts, with the aim of improving the quality and trustworthiness of published research in line with norms of reciprocity to the research community.” The eight pillars characterize manuscript peer review and are as follows: Principled, Constructive, Timely, Transparent, Professional, Quality, Confidential, and Pluralistic.

**Conclusions:** This three-round international Delphi established a consensus-based, normative conceptualization of manuscript peer review as a distinct, human-led process to improve the quality and trustworthiness of scholarly research. The resulting definition and eight pillars may provide a shared foundation for research, evaluation, education, and improvement across publishing contexts while allowing operational features to remain adaptable.

## Introduction

Peer review is widely recognized as a cornerstone of modern scientific publishing. Although its origins are often traced to the founding of the Royal Society’s Philosophical Transactions in 1665, practices resembling contemporary peer review developed gradually over subsequent centuries and varied across publication settings^1,2^. This historical variability, together with continuing differences in how peer review is conceptualized and implemented, underscores the need for a shared contemporary definition.

Today, peer review remains the primary mechanism for assessing methodological rigor and determining publication quality^2^. Despite this, calls for its improvement have intensified as multiple shortcomings in the current system have been identified^3,4^.

Rigorous evaluation of peer review practices, such as through randomized trials and systematic reviews, is needed to establish whether peer review truly improves research quality. One such analysis, a systematic review by Bruce and colleagues^5^ found significant variability in how peer review is conceptualized, concluding that an operational consensus definition is an important first step toward advancing such research on this topic. These authors also identified substantial conceptual variation and called for an operational consensus definition as a necessary step toward rigorous research to improve manuscript evaluation. Without definitional clarity, studies may evaluate fundamentally different processes under the same label.

This need for definitional clarity extends beyond research on peer review itself. In the presence of questionable publishing practices, including paper mills and predatory journals^3,6^, definitional clarity for peer review is increasingly important. Research integrity depends on transparency, ethical oversight, and credible peer review^7,8^. Consequently, understanding what constitutes peer review and, more importantly, what doesn’t is necessary for researchers, clinicians, and policymakers to distinguish legitimate practices from flawed or deceptive ones. Establishing this clarity is important for safeguarding the credibility of scientific publishing more broadly.

Although a clear and internationally applicable definition of manuscript peer review is needed to support research and practice, an international consensus definition of manuscript peer review has not been developed, contrasting sharply with many clinical domains. Several considerations may help explain the difficulty of developing a universally applicable definition, including the variation of peer review practices across disciplines^3^, the coexistence of multiple review models (e.g., single-blind, double-blind, post-publication, collaborative)^2^, and the rapidly evolving science dissemination landscape, including the use of preprints^9^ and movement towards increased open science and transparency practices^10^. Taken together, these challenges highlight the need to establish a shared conceptual foundation to support research and practice.

This study represents the second and final stage of a research program to establish an international consensus definition of manuscript peer review. The first stage involved a scoping review^11^, which identified definitions and characteristics of manuscript peer review in the literature. This second stage is a modified Delphi consensus study. Delphi studies structure communication among participants through multiple rounds of surveys, with aggregated responses presented between rounds to facilitate consideration of the group’s views and reassessment of individual responses^12–14^. The outcome of this study is an internationally agreed definition of manuscript peer review. Such a definition is expected to constitute a critical foundation for future research, systematic evaluation, and improvements to the scientific process.

### Study Aim

To conduct a Delphi process to inform an international, consensus definition of manuscript peer review.

## Methods

### Approach

A similar methodology used to achieve an international consensus definition of predatory publishing^6,15,16^ was used. A three-round modified Delphi was conducted (Rounds 1 and 2 as online surveys; Round 3 as a virtual consensus meeting). In the current context, the scope of the definition is focused on the review of manuscripts, including both pre-publication and post-publication review (e.g., eLife^17^). A ‘manuscript’ refers to the primary written draft of a scholarly work submitted for peer review. This scope extends to original research articles and conference proceedings but excludes opinion pieces such as editorials. Beyond establishing an operationally useful description of manuscript peer review, the definition is intended to be normative, identifying the values, purposes, and standards that *ought* to characterize the process, similar to Merton’s normative conception of science^18^.

### Ethics Approval and Transparency Practices

Ethics approval was obtained by the Ottawa Hospital Research Ethics Board (ID 20260027-01H). The study protocol was registered on the Open Science Framework (OSF)^19^ on May 13, 2026 at https://osf.io/z597r/files/ft2zx^20^. Study materials are available at: https://osf.io/w5gy8/files/osfstorage. This paper has been posted as a preprint on MedRxiv^21^ at: [insert link]. The Accurate Consensus Reporting Document (ACCORD) checklist^22^ and Method Reporting with Initials for Transparency (MeRIT)^23^ have been used to report findings (see **Table S1**).

Potential participants were informed that submission of the questionnaire constituted implied consent in Rounds 1 and 2. Round 3 participants provided consent through an electronic, written informed consent form on Microsoft Forms. For Rounds 1 and 2, all survey responses were de-identified before being analysed. Participants were assigned a unique ID to link survey responses, with contact information and corresponding IDs stored separately from survey data in a secure master list. For Round 3, voting was conducted anonymously. Participant names within the consensus meeting transcript record were replaced with unique participant IDs, and a separate master list was used to link participant IDs to individual participants. The master lists from all rounds, as well as the video recording from Round 3, were securely stored on the TOH/OHRI (The Ottawa Hospital/Ottawa Hospital Research Institute) server, which is password protected. Access was restricted to authorized members of the study team.

### Steering Committee

The steering committee comprised MMa, KHT, JSR, JT, MMi, CVR, JM, AB, IB, JC, MO, LW, and DM. The committee brought together individuals with diverse expertise and perspectives, including three patient partners (JT, MMi, and CVR). The steering committee provided overall oversight and strategic guidance throughout the study, including the development and refinement of survey items across all three Delphi rounds, methodological decision-making, and interpretation and dissemination of the findings. The patient partners contributed as full members of the steering committee, helping to ensure that patient-centered perspectives were incorporated throughout the study. Steering committee members were not eligible to participate in any Delphi round, including item voting. In addition to the steering committee, the Round 3 consensus panel included participants with expertise in patient and public involvement and engagement in research, bringing stakeholder-informed perspectives to the final deliberations.

### Scoping Review and Generation of Initial Survey Items

For Round 1, an initial list of items related to manuscript peer review was developed through a scoping review^11^. This review identified 4,181 sources that included definitions or characteristics of peer review, of which 139 contained empirically based definitions or characteristics. Thematic analyses of these 139 sources resulted in the creation of 37 peer review features that were classified under six themes (*Characteristics, Function, Motivation, Assessment, Process,* and *Challenges*). The research team (KHT, MMa, DM) reviewed and refined these features by separating multi-component items and simplifying wording to improve clarity, for example, by revising terminology relating to non–English language speakers. This process resulted in 45 candidate items. Steering committee members were also invited to propose additional items not identified in the scoping review. One additional item under the *Function* theme (‘peer review provides patient and public partner peer reviewers with recognition and credibility’) was added, resulting in 46 items for the Round 1 survey. Participants in all rounds were informed that the items had been developed from an initial scoping review. The preprint reporting the scoping review was provided to participants in Rounds 2 and 3 to support their consideration of the items during subsequent rounds.

### Participants and Recruiting

Participants were purposively recruited from a diverse group of international experts that represent interest holders (i.e., interest holders^24^) involved in manuscript peer review within the fields of biomedical sciences (e.g., molecular biology, genetics, and pharmacology), health sciences (e.g., medicine, nursing, public health, and allied health), social sciences (e.g., psychology, sociology, political science, and law), economics, and healthcare-related industries (e.g., pharmaceutical, diagnostic, and biotechnology sectors). Respondents were required to speak and read English to participate. To be included in the analysis of a given round and to remain eligible for the subsequent Delphi round, Round 1 and 2 participants were required to complete all survey items, excluding open-text questions. A “prefer not to say” option was available for all closed-text demographic questions. Participants did not receive remuneration or reimbursement for participation; however, authorship was offered to individuals who completed all three rounds. **Figure 1** illustrates participant progression across Delphi rounds.

**Figure 1.**
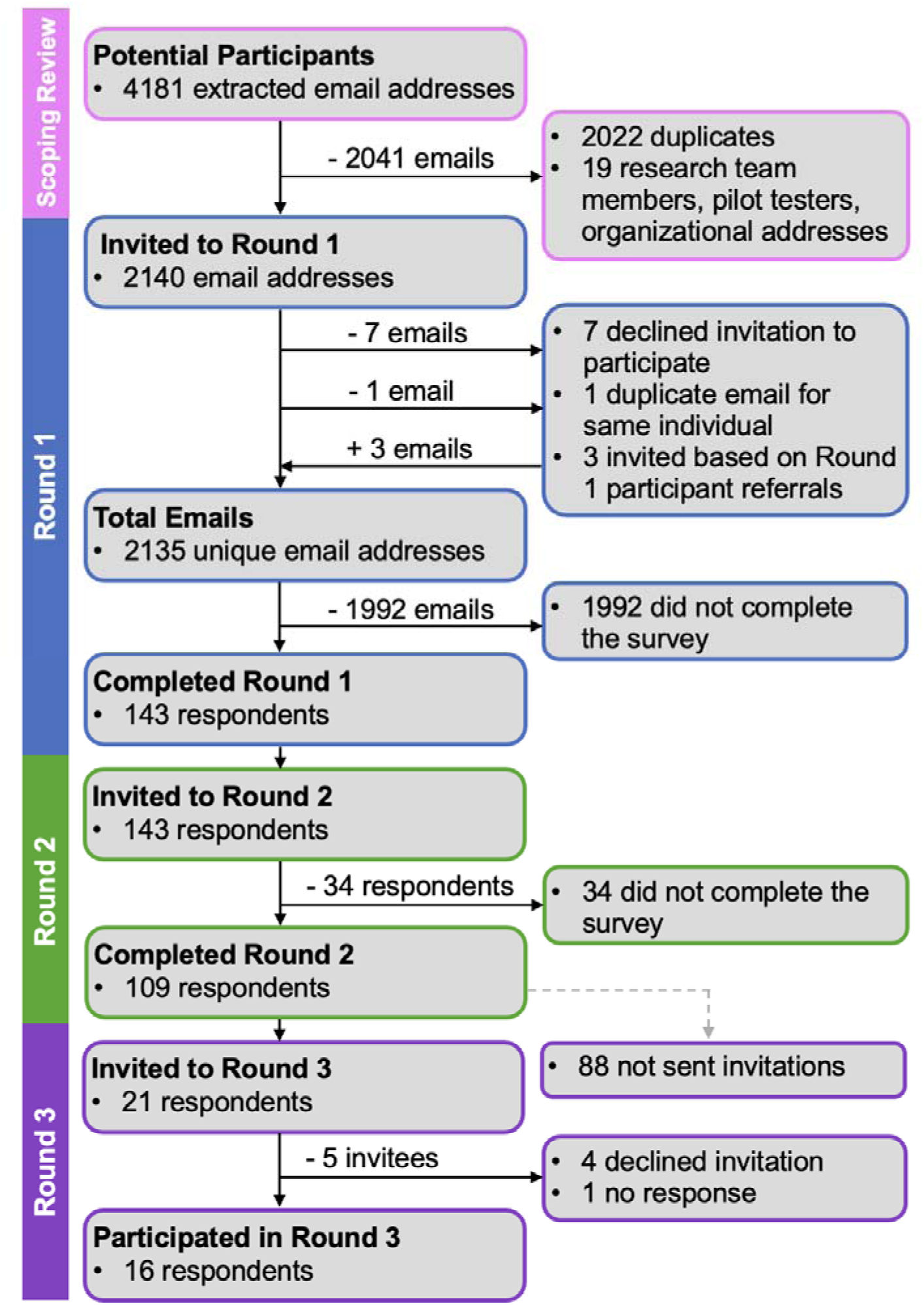
Participants Flowchart.

Round 1 invitees were corresponding authors from all articles included in the aforementioned scoping review^11^. Of 4,181 extracted email addresses, 2,022 duplicate addresses and 19 addresses associated with research team members, pilot testers, or organizations were excluded, yielding 2,140 initial invitees. Following the removal of seven individuals who declined participation and one duplicate individual entry, along with the addition of three individuals recommended by Round 1 participants, 2,135 eligible invitees remained, of whom 143 completed the survey. All Round 1 participants were invited to Round 2 (n = 143), of whom 109 completed the survey (109/143, 76.2% response rate).

Round 3 invitees were selected by the steering committee from Round 2 completers to address gaps in representation from earlier rounds and to broaden diversity of perspectives. Selection considered geographic location, gender/sex, race/ethnicity, age, professional role, career stage, field, and peer-review experience, informed by an equity lens and the PROGRESS-Plus framework^25,26^. To facilitate this process, research team members were provided with the following participant demographic information: full name, sex/gender, race/ethnicity, age, country of work, role (e.g., student, researcher, publisher), institution, career stage, primary field of research or professional practice, and experience with peer review. Non-demographic survey responses were not linked to these data. Selected participants received email invitations containing study information and an electronic informed consent form (Microsoft Forms). Invitations were sent between July 6 to July 16, 2026, with up to two weeks provided for response and follow-up at one week when applicable. Of the 21 invitations sent, 16 participants accepted and participated in Round 3 (BA, OBA, HB, KBM, GC, AF, DAF, ABK, MSK, GL, GM, DPM, ABRS, LSB, AKT, SY).

### Delphi Administration

#### Rounds 1 and 2

The first two Delphi rounds were online surveys administered using Welphi^27^. Participants received a standardized recruitment email containing a unique Welphi survey link that provided access to the implied consent form followed by the survey. Each round was designed for completion within 30 minutes. Round 1 was administered between April 13 to May 1, 2026, with one reminder being sent on April 22, 2026. Round 2 was administered from June 30 to July 15, 2026, with reminders sent on July 7, 2026 to non-responders and July 13, 2026 to participants who had initiated but not completed the survey.

Both surveys were pilot tested by individuals external to the steering committee to assess technological functionality and item comprehensibility. Feedback improved clarity without altering item content. Nine individuals pilot tested Round 1 between March 29 and April 7, 2026, and three individuals tested Round 2 between June 24 and June 26, 2026. Pilot testers were excluded from study participation.

In Round 1, a survey (including the ‘introduction’ and ‘informed consent form’ pages) was administered (see **Appendix A** for full list of survey items). A total of 92 survey questions were included under the following domains: (1) participant demographics and respondent peer review practices (13 questions); (2) peer review terminology and consensus development (22 questions); (3) peer review features (55 questions, including 46 items on peer review features generated from the previously described scoping review and grouped under the themes of *Characteristics, Function, Motivation, Assessment, Process,* and *Challenges*); and (4) additional comments and conclusion (2 questions). In the ‘peer review features’ category, participants rated the importance of 46 peer review features on a 5-point Likert scale [(1) Strongly disagree, (2) Disagree, (3) Neutral, (4) Agree, (5) Strongly agree]. The 5-point Likert scale was selected based on recommendations by previous studies^28,29^. Using free text boxes, participants were also asked for any additional comments on the voting items. Following administration, de-identified results from Round 1 were synthesized and analysed. The steering committee was then given an opportunity to modify survey items.

The same procedure used in Round 1 was used for Round 2. Participants were provided with a 66-item survey administered across 21 survey pages (including the ‘introduction’ and ‘informed consent form’ pages; see **Appendix B** for the full list of survey items). The survey included the following sections: (1) peer review terminology and consensus development (21 questions); (2) peer review features (42 questions, including 33 items on peer review features grouped under the themes of *Characteristics, Function, Motivation, Assessment, Process,* and *Challenges*); (3) demographics (1 question); and (4) additional comments and conclusion (2 questions). Similar to Round 1, respondents were asked to rate the importance of 33 items within the ‘peer review features’ section using the same 5-point Likert scale, with text boxes being used to solicit additional comments. Participants were also provided summarized, quantitative responses for items that did not reach consensus following Round 1. Responses informed steering committee survey item modifications. Demographic information was not collected in Round 2, with the exception of participants’ sex, which had been inadvertently omitted from Round 1. All other demographic characteristics reported for Round 2, including participant peer review practices, are based on Round 1 responses.

#### Round 3 (Online Summit)

A virtual summit was conducted through Zoom^30^ on August 5, 2026. The summit was moderated by DM, a researcher experienced with Delphi studies and consensus processes, and MMa. The Zoom^30^ platform was used to video-record the meeting, and to generate a verbatim transcription record of the round, which was subsequently de-identified. A note-taker from the Methodological and Implementation Research Program at OHRI documented key discussion points, while two additional program members supported slide management and vote tallying. None of these individuals were members of the steering committee. Three technology test runs were conducted before the meeting. For advance preparation, a booklet containing meeting instructions, items reaching consensus in Rounds 1 and 2, and all voting items was provided to participants on July 29, 2026 (see **Appendix C**).

The summit began with a summary of Round 1 and 2 results and items reaching consensus. Participants then discussed and voted on the workflow that best represents peer review based on Round 2 results, followed by a vote with the following options: (i) “*Separate Stage*” (i.e., peer review is the scholarly assessment of the manuscript; editorial decisions and publication are separate stages); (ii) “*Integrated Workflow*” (i.e., peer review encompasses the entire workflow, including reviewer assessment, editorial decisions, and publication); or (iii) “abstain from vote”.

This section of the meeting was moderated by AF, a participant that took part and voted during the consensus meeting with extensive expertise in editorial practices.

The 23 items that did not reach consensus in Round 2 were presented for discussion and voting at the consensus meeting, with items being grouped by themes (i.e., *Characteristics, Function, Motivation, Assessment, Process,* and *Challenges*). After each discussion round, an anonymous electronic vote was held using Microsoft Forms with responses being recorded using the following options: (i) “include”, (ii) “exclude”, or (iii) “abstain from vote”. Participants were provided the opportunity to modify items based on the discussion prior to voting.

A prototype definition, developed from items reaching consensus in Rounds 1 and 2 and the proposed workflow boundary, was presented for discussion. Participants provided feedback on the prototype definition, which was overwhelmingly supportive; given the strong level of agreement, no further formal endorsement via vote was deemed necessary, and only minor refinements were made based on participant feedback. Following this, a focused discussion on how to best implement the proposed definition was facilitated by DM. French’s Framework^31^ for theory-informed behaviour change interventions was used to offer context. A brainstorming exercise structured around addressing the four steps of French’s Framework with regard to implementing the new definition was used: (1) ‘Who needs to do what, differently?’; (2) ‘Which barriers and enablers need to be addressed?’; (3) ‘Which intervention components could overcome the modifiable barriers and enhance the enablers?’; and (4) ‘How can behaviour changes be measured and understood?’. To promote broad uptake, external interest holders (e.g., journal editors, publishers, funders, and institutional representatives) were recruited through knowledge users on the steering committee, engaged throughout the process, and supported as ambassadors for disseminating the final definition.

Demographic information was not collected during the consensus meeting; Round 3 demographic characteristics were derived from Round 1 responses, except sex assigned at birth, which was obtained from Round 2. The *Reviewer Characteristics* section, originally scheduled for the meeting, was voted on afterward due to time constraints. Participants received a Microsoft Forms survey with the six items, which remained open from August 7 to August 14, 2026, and had the same voting options as the meeting [(i) “include”, (ii) “exclude”, or (iii) “abstain from vote”].

#### Defining Consensus

In Rounds 1 and 2, consensus was defined as at least 80% (rounded to the nearest integer) of respondents rating an item within the top third (scores 4–5; include) or the bottom third of the scale (scores 1–2; exclude). Items reaching consensus were excluded from subsequent rounds. Items receiving _≥_80% neutral ratings (score 3) were considered to have ambiguous support and were retained for further consideration. In addition, the steering committee reviewed quantitative ratings and qualitative feedback after each round and removed items that were deemed unsuitable for further consideration.

In Round 3, a similar inclusion threshold of 80% represented majority consensus for the following options: (i) “include”, (ii) “exclude”. Consensus percentages were calculated solely based only on participants who selected either “include” or “exclude”; participants who selected “abstain” were excluded from the denominator. If consensus was not reached by Round 3, the item was not considered for inclusion in the final definition.

### Data Analysis and Presentation

The data collected in each round of the Delphi process was analysed individually. Frequencies and percentages were used to summarize the number of participants completing each round and their demographic characteristics (**Table 1**). Likewise, quantitative voting items were summarized through frequencies and percentages (see **Appendix D** for the complete list of quantitative results).

**Table 1.**
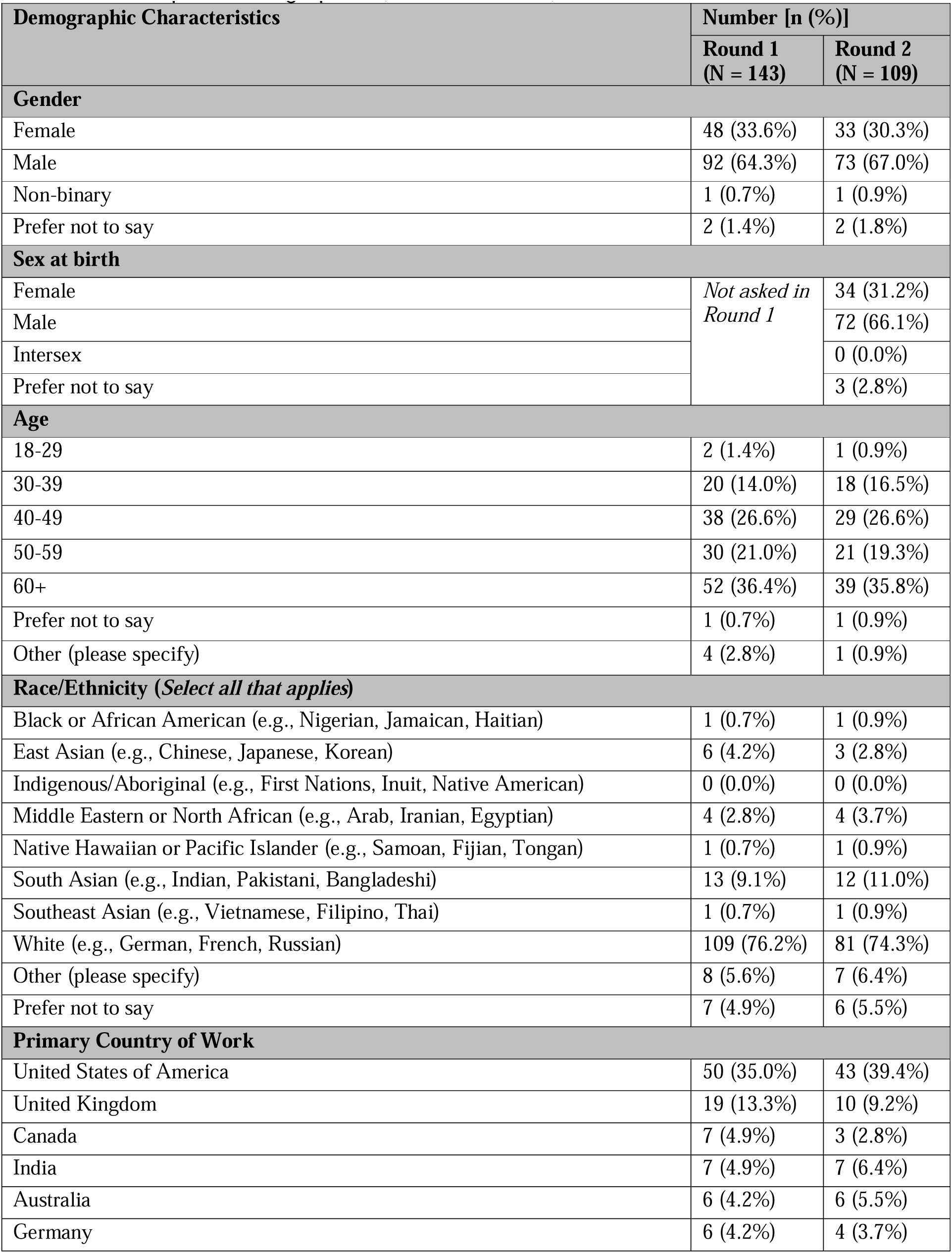
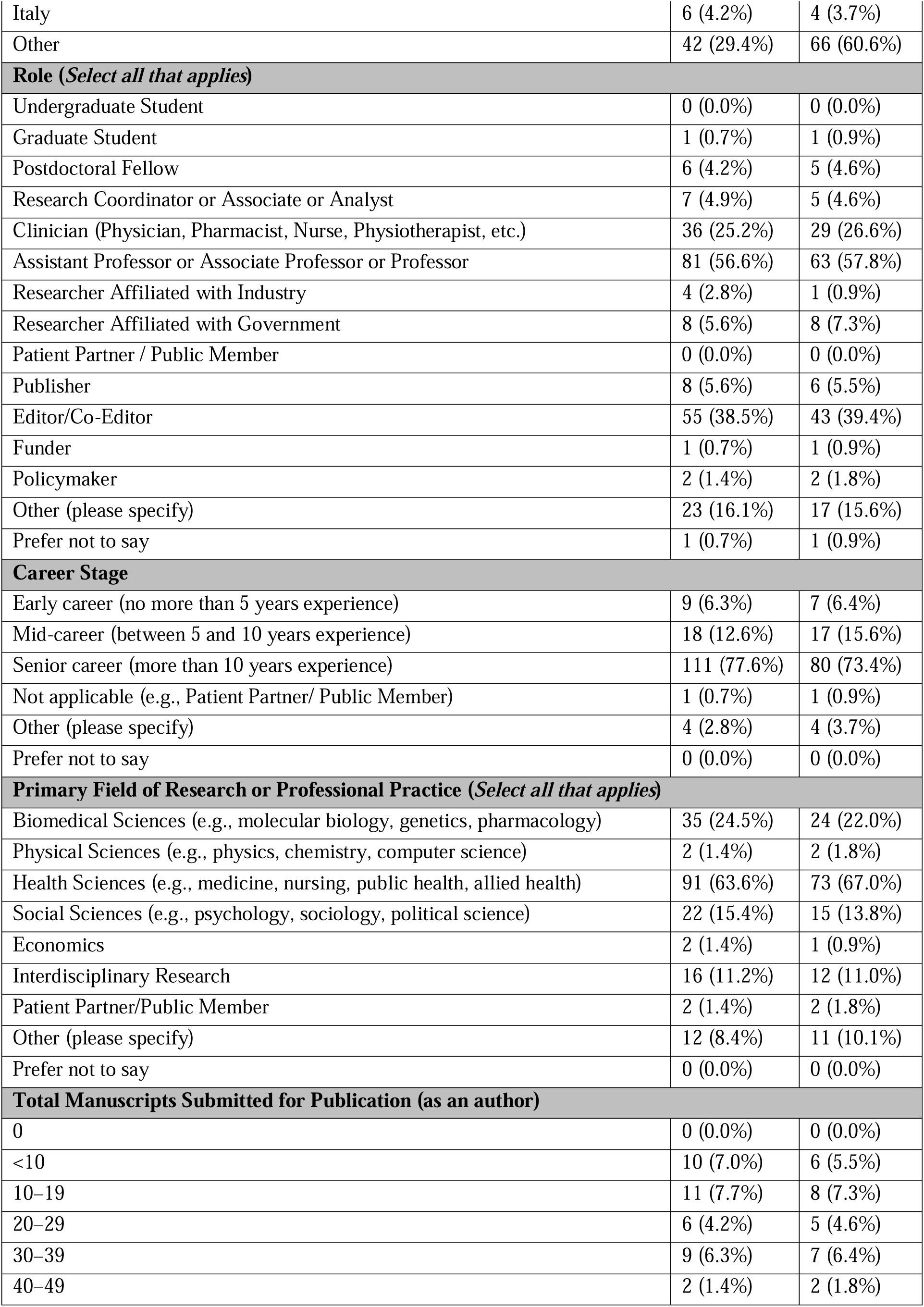
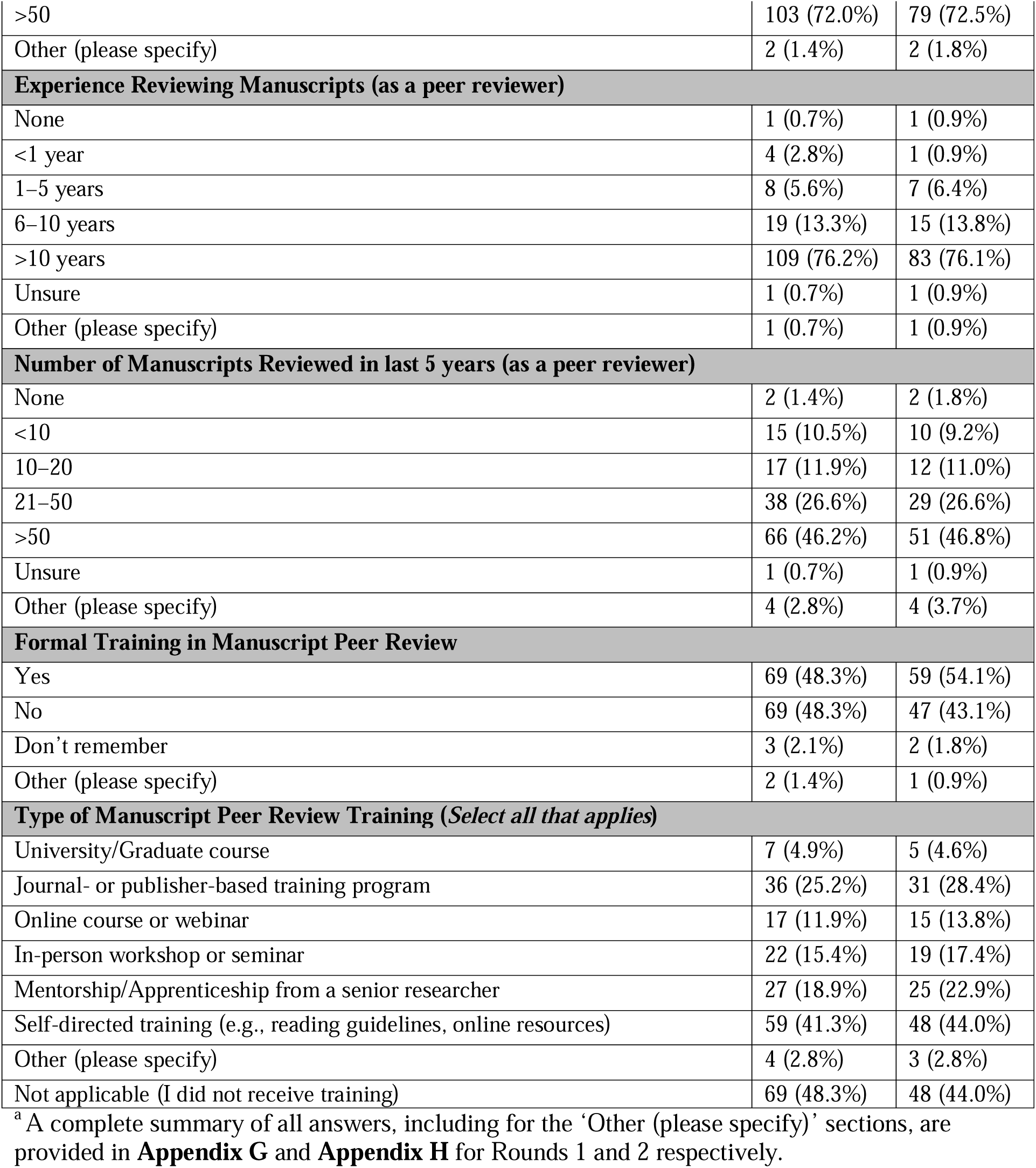
Participant Demographics (Rounds 1 and 2).

Qualitative, open-ended responses from the Round 1 and Round 2 surveys underwent an inductive analysis^32^. Generative artificial intelligence (ChatGPT^33^) was used to assist with the initial coding and thematic categorisation of free-text Delphi responses. All AI-generated codes and themes were subsequently reviewed, verified, and refined by (KHT, MMa, DM) through an iterative thematic analysis process. Three categories of information were analyzed: (1) item-specific responses on items that did not reach consensus; (2) additional comments and new item suggestions provided at the end of each survey section; and (3) responses to general open-ended questions about peer review (e.g., proposed solutions to current challenges facing peer review). Comments on items that met consensus were not analyzed. The analysis was conducted iteratively by KHT, with review by DM and MMa, and a code list was created for each qualitative question. Item-specific comments informed modification of existing items, while section-level comments informed development of new items. Responses to general open-ended questions informed additional clarification questions regarding the conceptualization of peer review. Full qualitative analyses are provided in **Appendix E** and **Appendix F** (for Rounds 1 and 2 respectively), with item progression summarized in **Figure 2** and specific modifications detailed in **Appendix D**.

**Figure 2.**
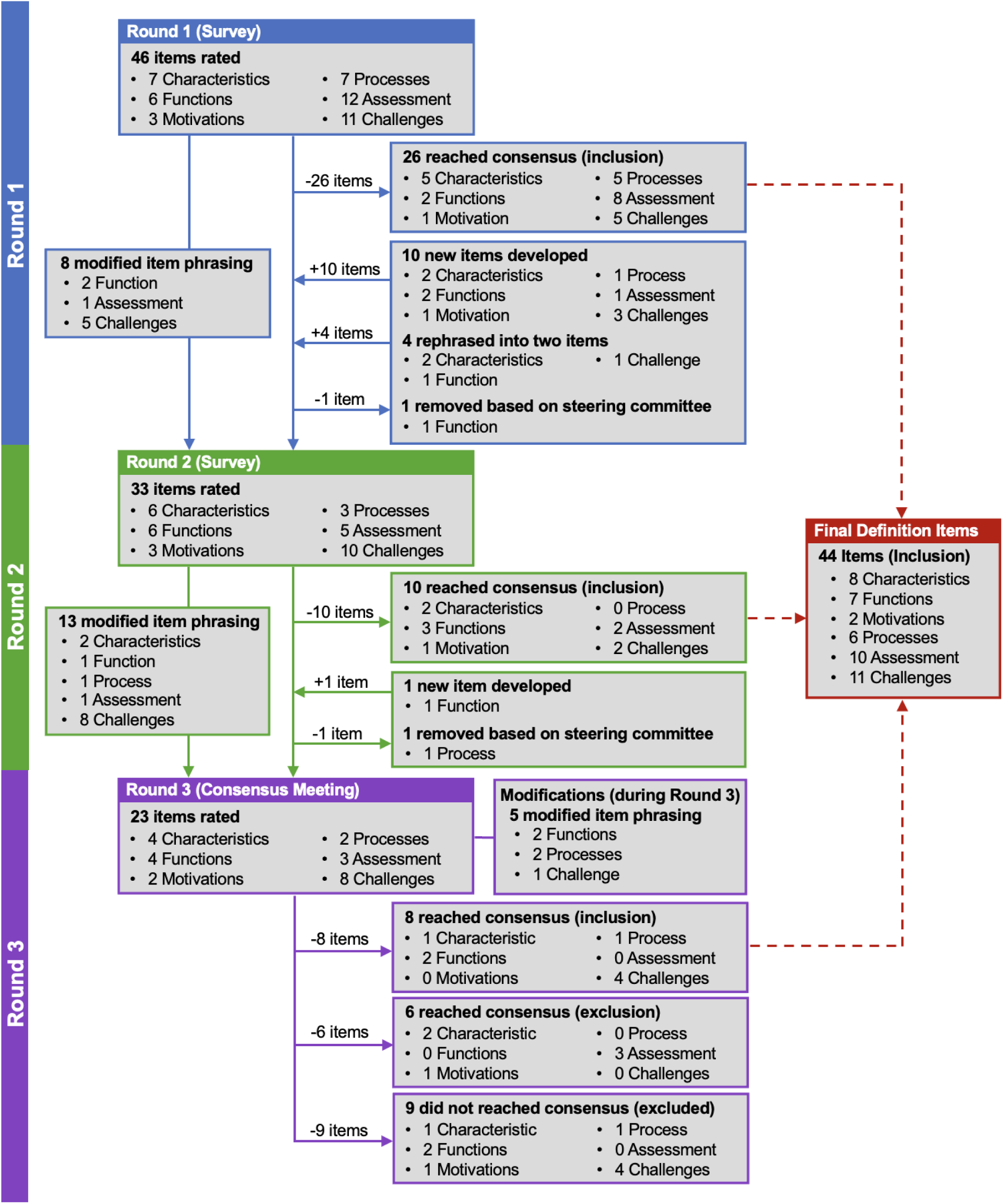
Peer Review Features: Item Tracking Flowchart.

#### Creating the Final Definition

An iterative process was used to develop the final definition. An initial prototype was developed from survey items reaching consensus by the end of Round 2, supplemented by relevant qualitative responses, and refined through steering committee discussion before presentation to Round 3 participants. The prototype was presented for discussion during Round 3 and received strong support, with no substantive concerns raised; participant feedback informed minor refinements to the wording. Following Round 3, consensus items and qualitative discussions from the summit were incorporated into the definition. The resulting manuscript, which provided the full definition and the complete explanation of each component (see [insert definition paper citation]), was circulated to Round 3 participants for feedback and further refinement before confirmation by the steering committee. The final conceptualization was derived from peer review features and reviewer characteristics that met consensus across the three rounds, the agreed terminology (“manuscript peer review”), the agreed conceptual boundary between peer review and editorial decision-making and publication, and discussion of the prototype definition during the consensus meeting.

## Results

### Deviations from the Protocol

Three deviations from the protocol^20^ occurred during the conduct of this study. First, participant recruitment was conducted exclusively through corresponding and, when relevant, first authors identified in the prior scoping review^11^. Second, the protocol originally specified that items would be removed solely based on predefined quantitative voting thresholds (_≥_80%). During the study, the steering committee also considered qualitative participant comments, alongside quantitative voting results, when making item removal decisions. Third, the consensus meeting was moderated by members of the steering committee (DM and MMa), rather than by an individual external to the steering committee as originally specified in the protocol.

### Round 1

A total of 143 participants completed Round 1. Of the remaining invited participants who did not complete the survey, only one progressed to approximately 50% of the survey; all others completed less than 10% (see **Appendix G** for the complete list of Round 1 responses).

#### Participants Demographics & Respondent Peer Review Practices

See **Table 1** for the complete list of demographic characteristics. Most respondents were male (gender; 92/143, 64.3%), White (109/143, 76.2%), over 50 years of age (82/143, 57.4%), and senior career professionals (>10 years experience; 111/143, 77.6%). Participants represented more than 120 institutions across over 35 countries, with the USA most represented (50/143, 35.0%). The most common professional roles were academic faculty (81/143, 56.6%), editors or co-editors (55/143, 38.4%), and clinicians (36/143, 25.7%), and health sciences was the predominant field (91/143, 63.6%). Most respondents had submitted more than 50 manuscripts for peer review in the past 5 years (103/143, 72.0%), had more than 10 years of reviewing experience (109/143, 76.2%), and had peer reviewed more than 50 manuscripts during this period (66/143, 46.2%). Fewer than half had received formal peer review training (69/143, 48.3%), most commonly through self-directed training (59/143, 41.3%) or journal-or publisher-based programs (36/143, 25.2%).

#### Peer Review Terminology & Developing Consensus

Among seven terms evaluated to describe peer review, ‘Manuscript peer review’ received the greatest support, with 108 participants (75.6%) rating it as a ‘good’ or ‘excellent’ name. Participants proposed more than 30 additional terms, which were synthesized into six options. These terms, together with ‘Manuscript peer review’, advanced to Round 2. When voting on the nine characteristics that reviewers should possess, four items met the criteria for consensus (two for inclusion; two for exclusion). Six items advanced to Round 2: five that did not reach consensus, all of which were rephrased, and one newly generated item. An additional survey question was also added based on participant responses to clarify if ‘peer’ was an appropriate term to describe reviewers within manuscript peer review.

A majority of the participants rated the development of a consensus definition as ‘very’ or ‘extremely’ important (82/143, 57.4%), while 43/143 (30.1%) considered it ‘moderately’ important. Nearly half reported that ‘some’ consensus currently exists regarding the definition of manuscript peer review (69/143, 48.3%), while none indicated that consensus exists across the entire community. Responses regarding established or consensus definitions were grouped into three categories: (1) conceptual sources identifying definitional gaps; (2) empirical studies examining the conduct and consistency of peer review; and (3) reform-oriented literature describing characteristics of stronger review systems. Several items explicitly point to definitional ambiguity, suggesting that the literature still lacks a universally accepted operational definition of manuscript peer review.

#### Peer Review Features

Of the 46 peer review features voted on, 26 reached consensus for inclusion and were removed from further voting. Following Round 1, four items were split into two items each, 10 new items were developed from participant responses, one item was removed following steering committee review of voting results and qualitative comments, and eight items were rephrased. These revisions resulted in 33 items grouped under the same six themes (i.e., *Characteristics, Function, Motivation, Assessment, Process,* and *Challenges*), which proceeded to Round 2 voting. See **Figure 2** for an overview of item changes.

Under the *Characteristics* section, over half of participants (78/143, 54.5%) indicated that some aspects of manuscript peer review may vary by context and therefore may not be universally applicable, including anonymity, confidentiality, and peer review expectations and objectives. Within the *Assessment* section, uncertainty emerged regarding whether editorial and publication decision-making, for example, should a manuscript be accepted for publication, should be considered within the scope of manuscript peer review. An additional Round 2 question was added to clarify this relationship. Under the *Challenges* section, respondents highlighted the importance of greater transparency, improved recognition and reward systems for reviewers, and enhanced editorial oversight as potential strategies to address challenges in manuscript peer review. Finally, when prompted about additional comments on developing a consensus definition, participants highlighted that consensus may be difficult to achieve across fields and cautioned that consensus does not necessarily equate to objective validity.

### Round 2

A total of 109 participants completed Round 2 (76.2% response rate; see **Table 1** for demographic characteristics). The complete list of Round 2 responses is provided in **Appendix H**.

#### Participants Demographics & Respondent Peer Review Practices

Round 2 participants were highly comparable to those in Round 1 with respect to demographic, professional, and peer review characteristics (see **Table 1**). The sample remained predominantly male - gender (73/109, 67.0%); sex assigned at birth (72/109, 66.1%)], White (81/109, 74.3%), over 50 years of age (60/109, 55.1%), and senior-career professionals (>10 years experience; 80/109, 73.4%). Participants represented more than 100 institutions across 30 countries and reported extensive peer review experience, with most having more than 10 years of reviewing experience (83/109, 76.1%) and having submitted more than 50 manuscripts for peer review in the past 5 years (79/109, 72.5%).

#### Peer Review Terminology & Developing Consensus

Similar to Round 2, the perceived importance of developing a consensus definition remained high, with 90/109 participants (82.6%) rating it as moderately, very, or extremely important. Of the seven terms assessed, ‘Manuscript peer review’ received the highest support, with 87/109 participants (79.9%) rating it as a good or excellent name, and was retained as the final terminology. Accordingly, terminology was not subject to further discussion or voting in Round 3. Consensus was reached to retain the term ‘peer’ for reviewers, with 88/109 respondents (80.7%) supporting its continued use. Qualitative feedback supported these results, citing its established use and capacity to encompass diverse expertise, perspectives, and backgrounds. Of the six *Reviewer Characteristics* voting items, two met the inclusion consensus threshold and were removed from subsequent voting; three new items were proposed based on qualitative feedback, and one was removed by the steering committee, leaving six items for Round 3.

When asked whether editorial and publication decision-making processes should be considered part of manuscript peer review, 60 respondents (55.0%) considered editorial processes separate from manuscript appraisal, while 46 (42.2%) supported their inclusion. Qualitative analysis of participant comments identified two perspectives: (1) *Separate stage*, in which editorial decision-making and publication occur independently of peer review; and (2) *Integrated workflow*, in which peer review encompasses reviewer assessment, editorial decisions, and publication. These frameworks were subsequently discussed and voted on during Round 3.

#### Peer Review Features

Following Round 2, a total of 10 out of the 33 peer review features achieved consensus for inclusion and were removed from further voting. Based on the Round 2 voting results and qualitative feedback provided by participants, the steering committee developed one new item, removed one item, and revised the phrasing of 13 items. These changes resulted in 23 items organized within the same six themes, which were subsequently advanced for voting during the consensus meeting. See **Figure 2** for an illustrated summary of item changes.

Within the Characteristics section, most respondents (70/109; 64.2%) agreed that characteristics of manuscript peer review may vary by context and therefore may not be universally applicable; for example, approaches to reviewer anonymity and confidentiality may differ across journals, disciplines, and peer-review models. Under the *Challenges* theme, respondents identified several potential solutions to address challenges in facilitating manuscript peer review, including the adoption of newer peer review models (e.g., registered reports and public participation in peer review), development of cross-journal reviewer registries, and maintenance of peer review records. Finally, when providing additional comments on the development of a consensus definition, participants emphasized the importance of ensuring that the definition allows for flexibility.

### Round 3

Round 3 invitees were purposely selected from Round 2 completers to address representation gaps and broaden the diversity of perspectives. Complete Round 3 voting results are provided in **Appendix I**.

#### Participants Demographics

Round 3 participants represented a diverse group in terms of demographic and professional characteristics. Gender was relatively balanced (male: 8/16, 50.0%; female: 6/16, 37.5%; non-binary: 1/16, 6.3%: 1/16, 6.3% did not answer), as was sex assigned at birth (female: 7/16, 43.8%; male: 7/16, 43.8%). Participants represented 12 countries across North America, South America, Europe, the Middle East, and Asia, with the largest age group being 40–49 years (7/16, 43.8%). Most participants were professors (11/16, 68.8%), editors or co-editors (9/16, 56.3%), and/or clinicians (5/16, 31.3%), were senior-career professionals (>10 years of experience; 10/16, 62.5%), and had a primary background in health sciences (13/16, 81.3%). See **Table S2** for a summary of Round 3 demographic information.

#### Peer Review Workflow, Features, & Reviewer Characteristics

Most participants supported conceptualising manuscript peer review as a separate stage from editorial decision-making and publication (13/15, 86.7%), rather than as an integrated workflow (2/15, 13.3%); one participant abstained. This conceptual boundary was retained in the final conceptualization. Of the 23 peer review feature items considered during the consensus meeting, five were rephrased following discussion. Fourteen achieved consensus (eight for inclusion and six for exclusion), whereas nine did not meet the consensus threshold. Figure 2 summarises item changes. Of the six Reviewer Characteristics items voted on, five reached consensus (two for inclusion and three for exclusion), while one did not meet the consensus threshold and was removed.

### The Final Definition

Following all three rounds, a total of 44 peer review features (i.e., *Characteristics, Function, Motivation, Assessment, Process,* and *Challenges*) and six reviewer characteristics met consensus for inclusion (see **Table 2** for the full list of items). The final conceptualization was informed by results from all rounds, including items voted on, qualitative comments from Rounds 1 and 2, and voting results on other aspects of peer review, such as terminology and workflow.

**Table 2.**
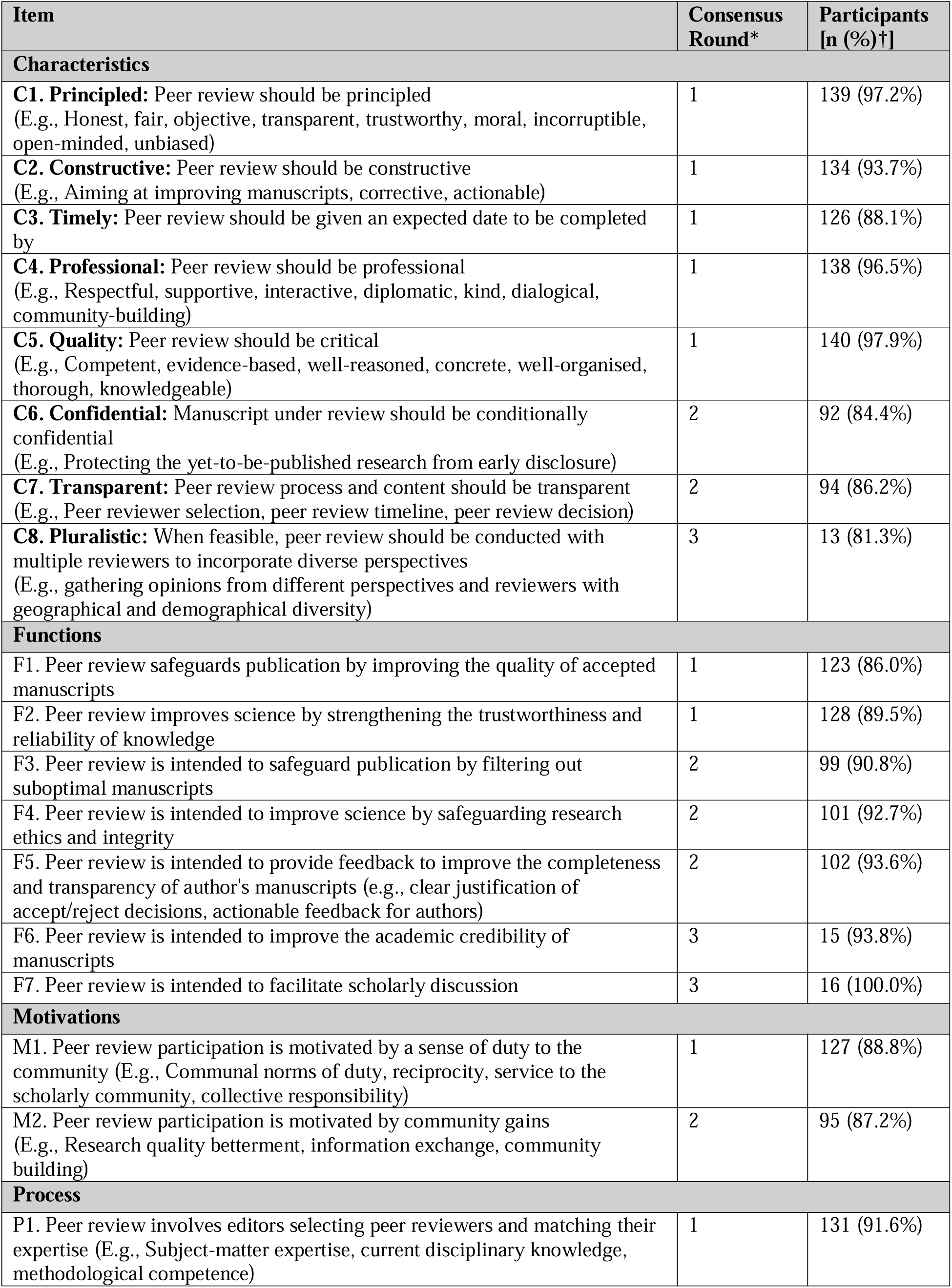
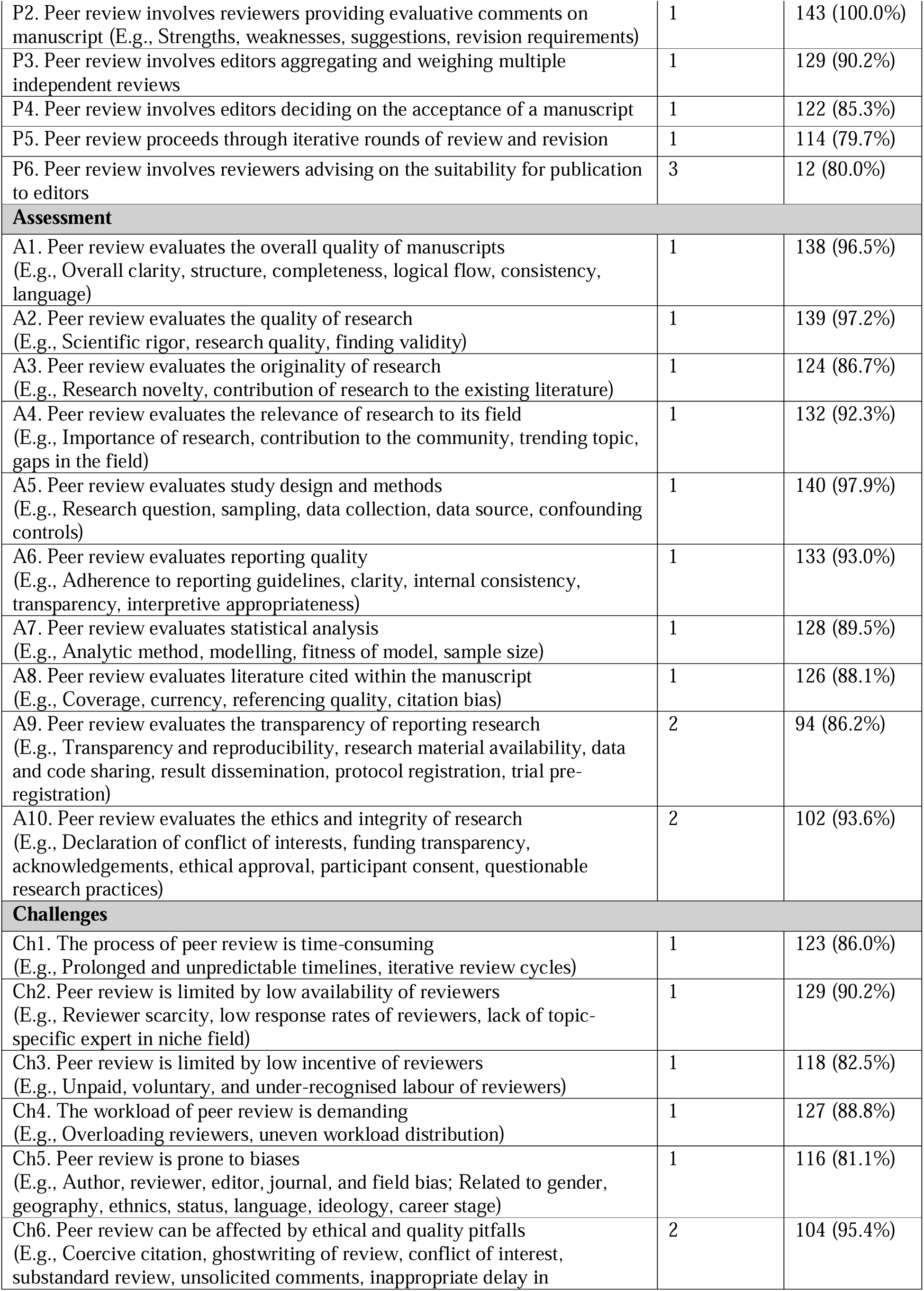
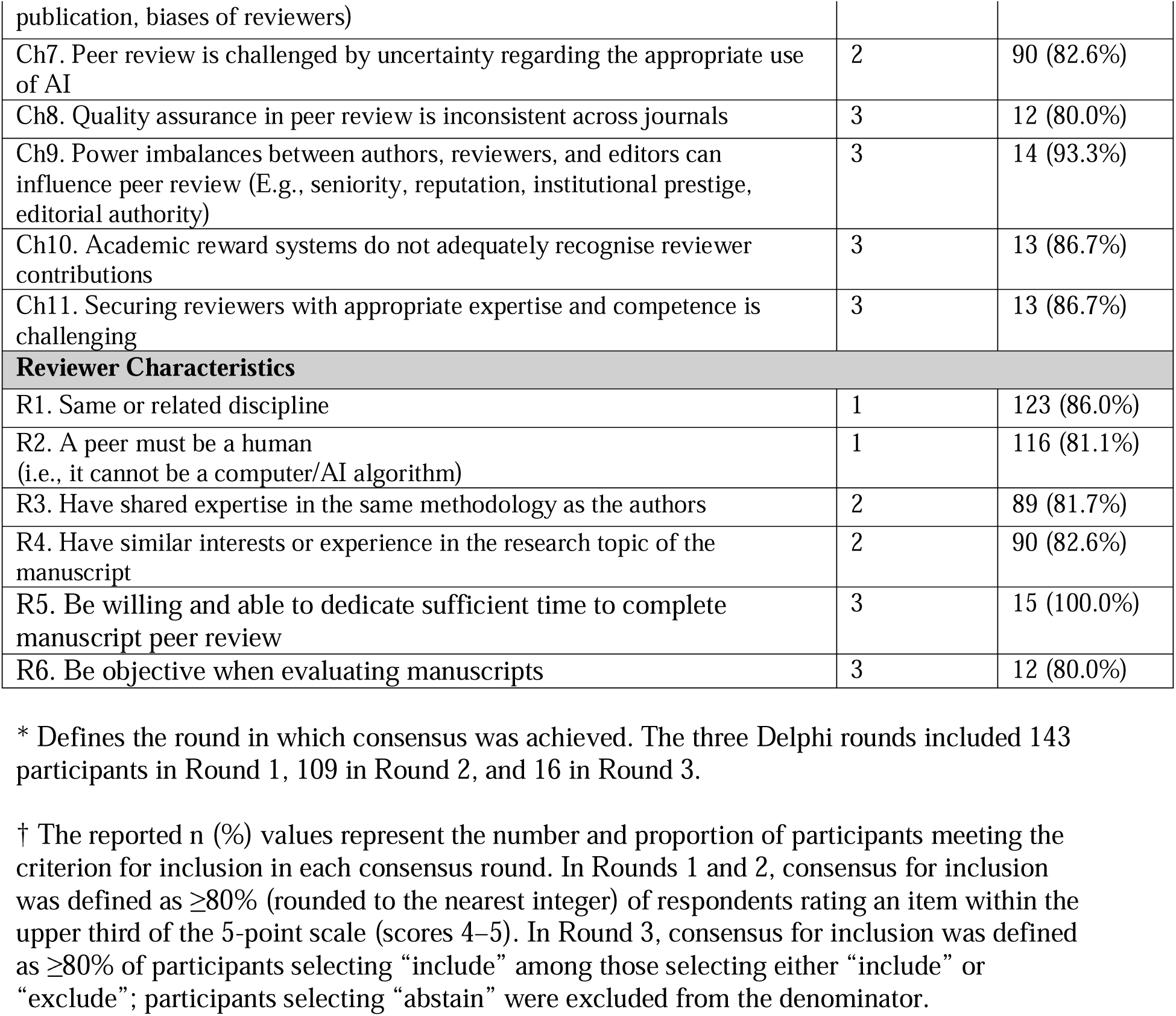
Consensus Items (Inclusion).

The final conceptualization of manuscript peer review comprises two components: a consensus definition and eight pillars. The consensus definition is: “Manuscript peer review is a structured process by which human reviewers assess scholarly manuscripts, with the aim of improving the quality and trustworthiness of published research in line with norms of reciprocity to the research community.” The eight pillars characterize manuscript peer review and are as follows: Principled, Constructive, Timely, Transparent, Professional, Quality, Confidential, and Pluralistic. A detailed description of this conceptualization, including an explanation of each component, is reported separately (cite definition manuscript).

## Discussion

This study aimed to develop a consensus definition of manuscript peer review. Across three Delphi rounds (with 143, 109, and 16 international participants, respectively), consensus for inclusion was reached on 44 features that characterize manuscript peer review and six reviewer characteristics, as well as on the preferred terminology and its position within the publication process. Participants endorsed ‘Manuscript peer review’ as the preferred term and defined it as a distinct stage of scholarly manuscript assessment. The resulting conceptualization highlights manuscript peer review as a structured, human-led process intended to improve the quality and trustworthiness of published research through reciprocity to the research community. It is underpinned by eight pillars (Principled, Constructive, Timely, Transparent, Professional, Quality, Confidential, and Pluralistic) and recognizes that some operational features should remain flexible across disciplines and publishing contexts.

Shared definitions provide a common language that supports consistency in clinical practice, research, and education. Clinical frameworks enable clinicians and researchers to classify comparable patients, apply evidence consistently, and meaningfully synthesize studies across settings, as, for example, in the definition of myocardial infarction^34^. An analogous consensus definition of manuscript peer review can provide a conceptual foundation for original research, meta-research, teaching, journal practices, and improvement efforts in scholarly publishing.

An international consensus definition of manuscript peer review establishes a common reference point for original research on the topic, enabling investigators to specify consistently what intervention or process they are studying. Studies have identified various roles or functions of peer review, for instance distinguishing between determining quality and providing legitimacy, or between certification, formation, allocation, and legitimation^35,36^. The definition of manuscript peer review resulting from our Delphi process aligns mostly with the formation function of peer review, which emphasizes the role of peer review in improving the information being reviewed.

For meta-research on manuscript peer review across disciplines, a consensus definition enables meaningful comparison, synthesis, and evaluation of peer-review models and outcomes across journals and disciplines. For teaching and training, it provides a clear foundation for educating authors, reviewers, editors, and patient and public partners about the process. For policy and practice, it provides a shared basis for developing, implementing, and evaluating standards and interventions related to manuscript peer review.

An important contribution of this work is that it establishes not only what manuscript peer review is, but also its conceptual boundaries within the broader publication pathway. Although Round 2 revealed differing views about whether editorial decision-making and publication should be considered part of peer review, the Round 3 panel reached consensus that manuscript peer review is a distinct stage of scholarly manuscript assessment, rather than the entire editorial and publication workflow (86.7% support). Under this conceptualization, editorial decision-making and publication are analytically distinct from manuscript peer review, although their timing and relationship to reviewer assessment may vary across publishing models, including pre-publication and post-publication review. This boundary is consequential: it enables researchers, journals, educators, and other interest holders to distinguish the effects and responsibilities of reviewer assessment from those of editorial judgement and publishing decisions, thereby supporting more precise reporting, measurement, comparison, and improvement of each component of the publication system.

### Strengths and Limitations

A strength of the Delphi methodology is its ability to facilitate input from all participants while reducing direct confrontation and the influence of power dynamics and hierarchies through the use of aggregated voting. Participant demographic and professional characteristics were also broadly similar across Rounds 1 and 2, supporting consistency in panel composition across survey rounds. The Round 3 consensus meeting included participants with diverse demographic and professional characteristics, including the Global South, contributing a broad range of perspectives to the consensus process and supporting the inclusivity of the resulting definition.

Although the conceptualization is intended to apply to both pre-publication and post-publication manuscript peer review, some consensus process features reflect common editor-mediated journal workflows. These features should be understood as operational elements that may vary across publishing contexts rather than as universal requirements of manuscript peer review. For example, post-publication, platform-based, and community-led models may organize reviewer selection, synthesis of feedback, and relationships with editorial decision-making differently while retaining the core purpose of scholarly manuscript assessment.

Several limitations should also be considered. First, although efforts were made to preserve nuances in language from the original scoping review^11^, some ideas may have not been fully captured, potentially influencing the phrasing and interpretation of survey questions. Second, requiring participants to speak and understand English limited the diversity of global perspectives represented. Finally, although efforts were made to recruit an internationally diverse panel, only a selected group of respondents contributed to the survey results^37^. Inclusion of a greater quantity of individuals with different expertise may have resulted in different study outcomes.

### Next Steps

This study addresses an important gap by providing an international, consensus-based definition and conceptualization of manuscript peer review^38,39^. However, a definition alone will not resolve the longstanding structural challenges of peer review; rather, it provides an essential foundation for studying the process more rigorously and comparing findings across settings. A logical next step is to establish consensus on which outcomes are used to measure manuscript peer review and to develop valid, feasible measures for assessing those outcomes. Such work would enable more informative evaluations of peer-review models and interventions, ultimately supporting evidence-informed improvements to peer review. Consequently, the Primary Evaluation outcomEs for Research on peer review – A Core Outcome Set (PEER-COS) project has been initiated and registered with the Core Outcome Measures in Effectiveness Trials (COMET) group^40^.

## Conclusions

Although developed for manuscript peer review, the definition may offer a useful starting point for conceptualizing peer review in other scholarly assessment contexts (e.g., the evaluation of researchers or research units), further work is needed to assess its applicability and potential need for adaptation in these settings.

This three-round international Delphi study established a consensus-based, normative conceptualization of manuscript peer review as a distinct, human-led process for improving the quality and trustworthiness of scholarly research through reciprocity to the research community. The definition and its eight pillars provide a common foundation for more consistent research, evaluation, education, and improvement of manuscript peer review across publishing contexts, while recognising that some operational features must remain adaptable. As a consensus-based normative framework, its applicability should be assessed across disciplines, languages, stakeholder groups, and diverse pre- and post-publication peer-review models to inform future refinement.

## Supporting information

Manuscript

ACCORD

## Data Availability

The study protocol was registered on the Open Science Framework (OSF)19 on May 13, 2026 at https://osf.io/z597r/files/ft2zx20. Study materials are available at: https://osf.io/w5gy8/files/osfstorage.

## List of Abbreviations

ACCORD: Accurate Consensus Reporting Document
COMET: Core Outcome Measures in Effectiveness Trials
MeRIT: Method Reporting with Initials for Transparency
OHRI: Ottawa Hospital Research Institute
PEER-COS: Primary Evaluation outcomEs for Research on peer review – A Core Outcome Set
PROGRESS-Plus: Place of residence, Race/ethnicity/culture/language, Occupation, Gender/sex, Religion, Education, Socio-economic status, Social capital
TOH: The Ottawa Hospital

## Declarations

### Ethics Approval and Consent to Participate

Research ethics approval was obtained by the Ottawa Health Science Network Research Ethics Board (ID 20260027-01H). Participants in Rounds 1 and 2 provided implied consent electronically on the Welphi platform. Participants in Round 3 provided consent through an electronic, written informed consent form on Microsoft Forms.

## Availability of Data and Materials

All relevant, de-identified data has been included in the final manuscript or posted on OSF (see https://osf.io/w5gy8/files/osfstorage).

## Competing Interests

JC is the International Editor of the BMJ. LW is Co-Editor-in-Chief of MetaROR. OBA is one of the coordinators of the Brazilian Reproducibility Network and is part of eLife’s Global South Committee for Open Science. HB is the Chair of Scientific Advisory Board, Medeloop Scientific Advisory, Explore Science Member, Scientific Advisory Board, Vivili. GC is Senior Strategy Advisor, Open Research at UKRI. He has published editorials advocating for open and post-publication peer review and previously worked at eLife where he promoted the publish-review-curate model. AF is Executive Managing Editor and VP Editorial Operations, JAMA and JAMA Network; Executive Director, International Congress on Peer Review and Scientific Publication; and serves on the Editorial Boards of Research Integrity and Peer Review, Nursing Outlook, and American Journal of Nursing. DAF is an editorial board member of several international journals (BMC Research Notes, PLOS One, American Journal of Medical Genetics B and WIREs Mechanisms of Disease). ABK was Executive Editor and Editor-in-Chief of the International Journal of Therapeutic Massage and Bodywork from 2016 to 2022 and has previously published editorials and scholarship concerning manuscript peer review and scientific publishing. She reports no financial competing interests related to this work. MSK is the Editor-in-Chief of Indian Journal of Anaesthesia, Associate Editor of Journal of Anaesthesiology Clinical Pharmacology, Associate Editor of Practical Evidence in Anaesthesia Knowledge.

GM has received honoraria from Sanofi® and AstraZeneca®. DPM is the Editor-in-Chief of the Indian Journal of Rheumatology, an Associate Editor at International Journal of Rheumatic Diseases, Frontiers in Immunology, Future Healthcare Journal, and Journal of Inflammation Research, and serves on the Editorial Board of Clinical Rheumatology, Mediterranean Journal of Rheumatology, Archives of Rheumatology, and Rheumatology and Therapy. SY is Associate Editor of Proceedings, The Official Journal of Shaikh Zayed Post Graduate Medical Institute. DM is a member of the BMJ’s North American advisory board and an editorial board member of the Journal of Clinical Epidemiology, BMC Medicine and subject editor for FACETS.

### Patient and Public Involvement

Three patient partners (JT, MMittelman, CVR) were part of the study team and were involved with the conduct and reporting of this study. Their engagement supported the integration of patient-centered perspectives into the interpretation and dissemination of findings. In addition to the steering committee, the Round 3 consensus panel included participants with expertise in patient and public involvement and engagement in research, bringing stakeholder-informed perspectives to the final deliberations.

## Acknowledgements

We deeply thank everyone for their assistance in this project. We acknowledge the following individuals for pilot testing Round 1: Cora Burgwinkel, Minoo Matbouriahi, Florian Naudet, Jeremy Y. Ng, Fiona Ramage, Nino Tsertsvadze, Giulia Varvarà, Constant Vinatier, and Jifan Wang. Similarly, we acknowledge the following individuals for pilot testing Round 2: Marc A. Albert, Laura Caquelin, Camille Rousseau. We acknowledge Monika Szejbach for booklet creation and note-taking, Daphne Eunice Ramírez García for slide management, and Anna Catharina Vieira Armond for vote tallying during the Round 3 consensus meeting.

## Online Appendices (OSF Uploads)

**Appendix A.** Round 1 Survey Questions. [https://osf.io/w5gy8/files/euzdk^41^]

**Appendix B.** Round 2 Survey Questions. [https://osf.io/w5gy8/files/hk936^42^]

**Appendix C.** Round 3 Booklet. [https://osf.io/w5gy8/files/he927^43^]

**Appendix D.** Quantitative Voting Item Results (All Rounds). [https://osf.io/w5gy8/files/kj2nv^44^]

**Appendix E.** Qualitative Analysis Sheet (Round 1). [https://osf.io/w5gy8/files/cuj48^45^]

**Appendix F.** Qualitative Analysis Sheet (Round 2). [https://osf.io/w5gy8/files/9pb7y^46^]

**Appendix G.** Round 1 Results (Complete Summarized). [https://osf.io/w5gy8/files/a8xuk^47^]

**Appendix H.** Round 2 Results (Complete Summarized). [https://osf.io/w5gy8/files/6xb3f^48^]

**Appendix I.** Round 3 Results (Complete Summarized). [https://osf.io/w5gy8/files/ec24m^49^]

## Supplementary Files

**Table S1.**
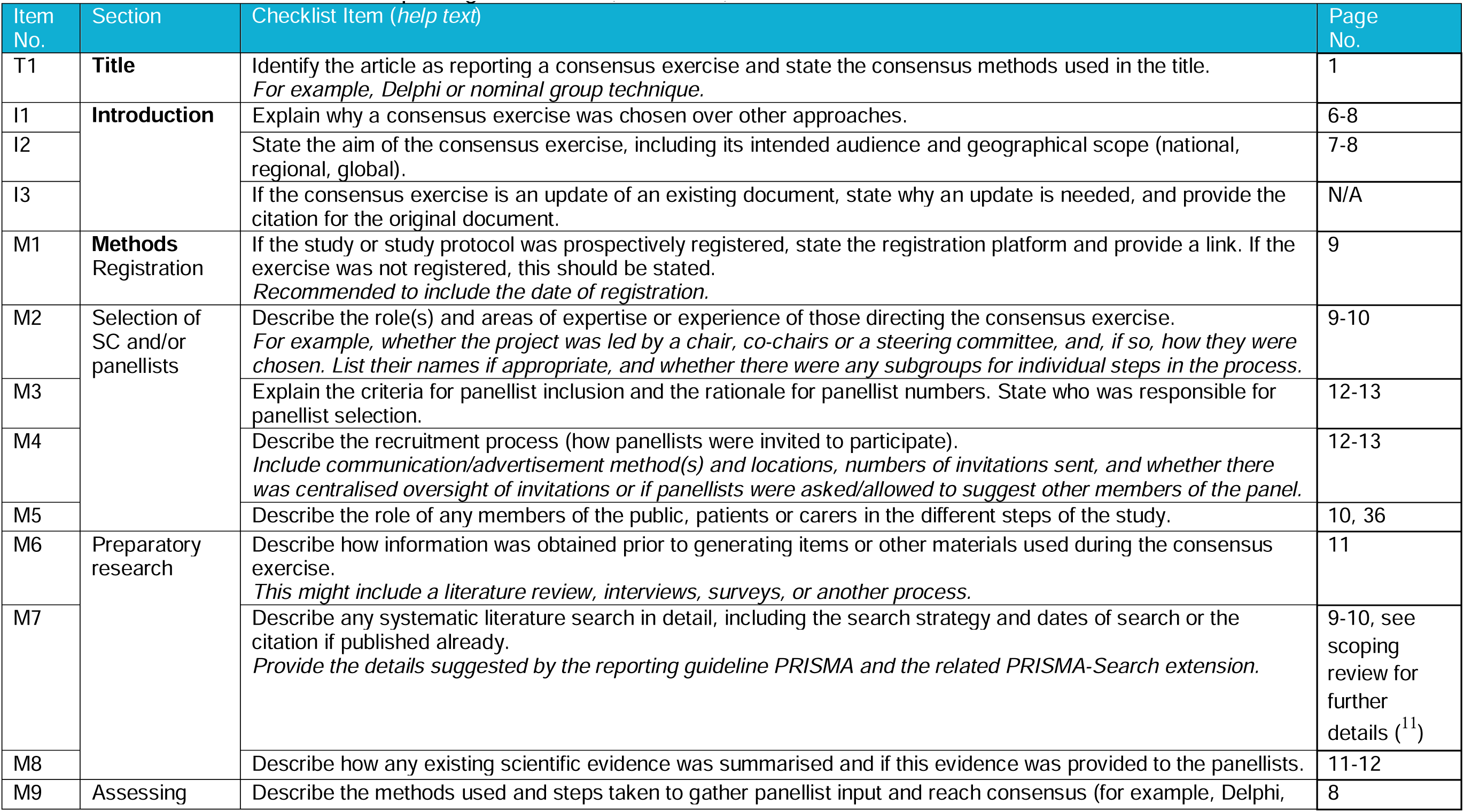
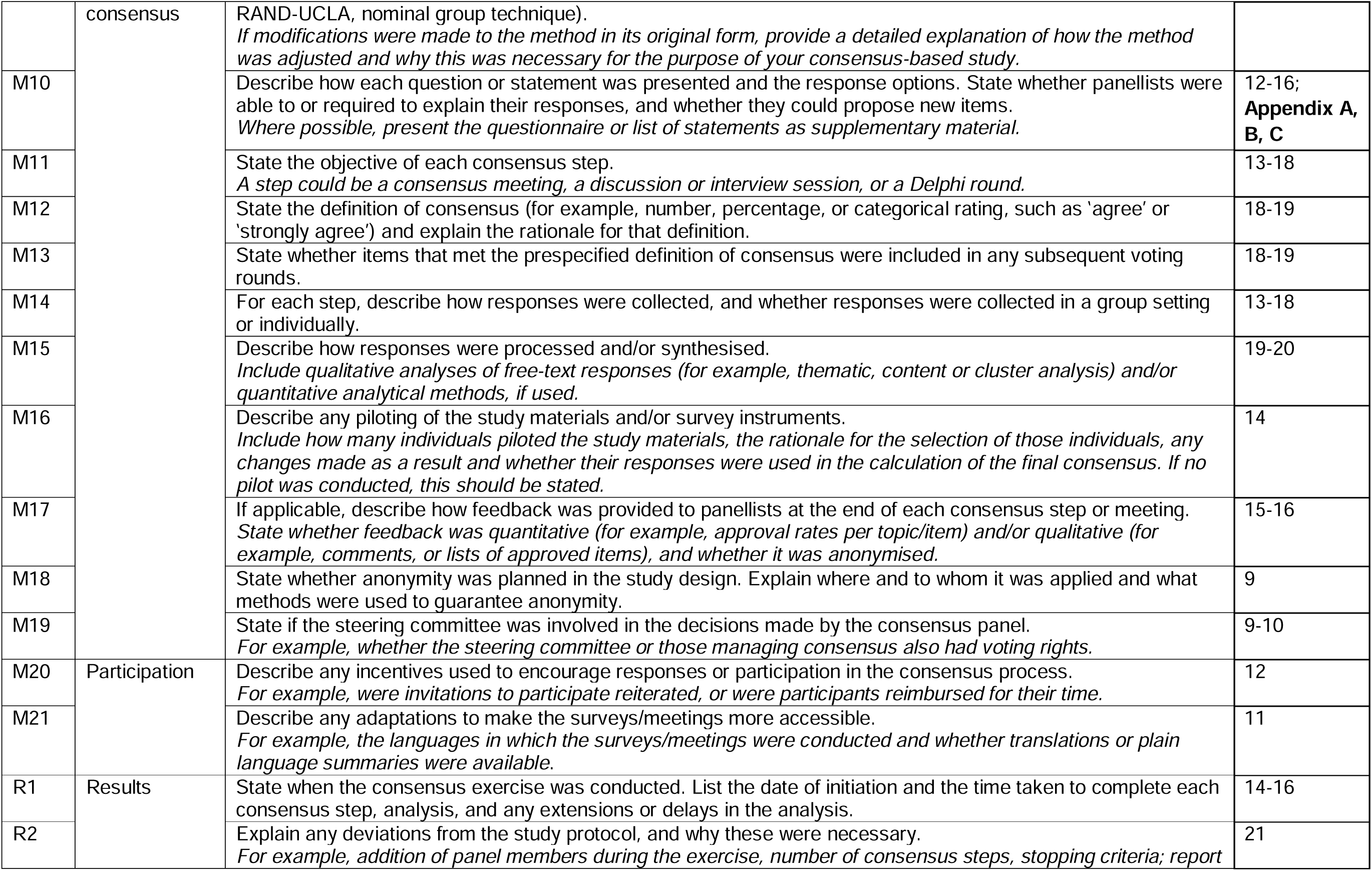
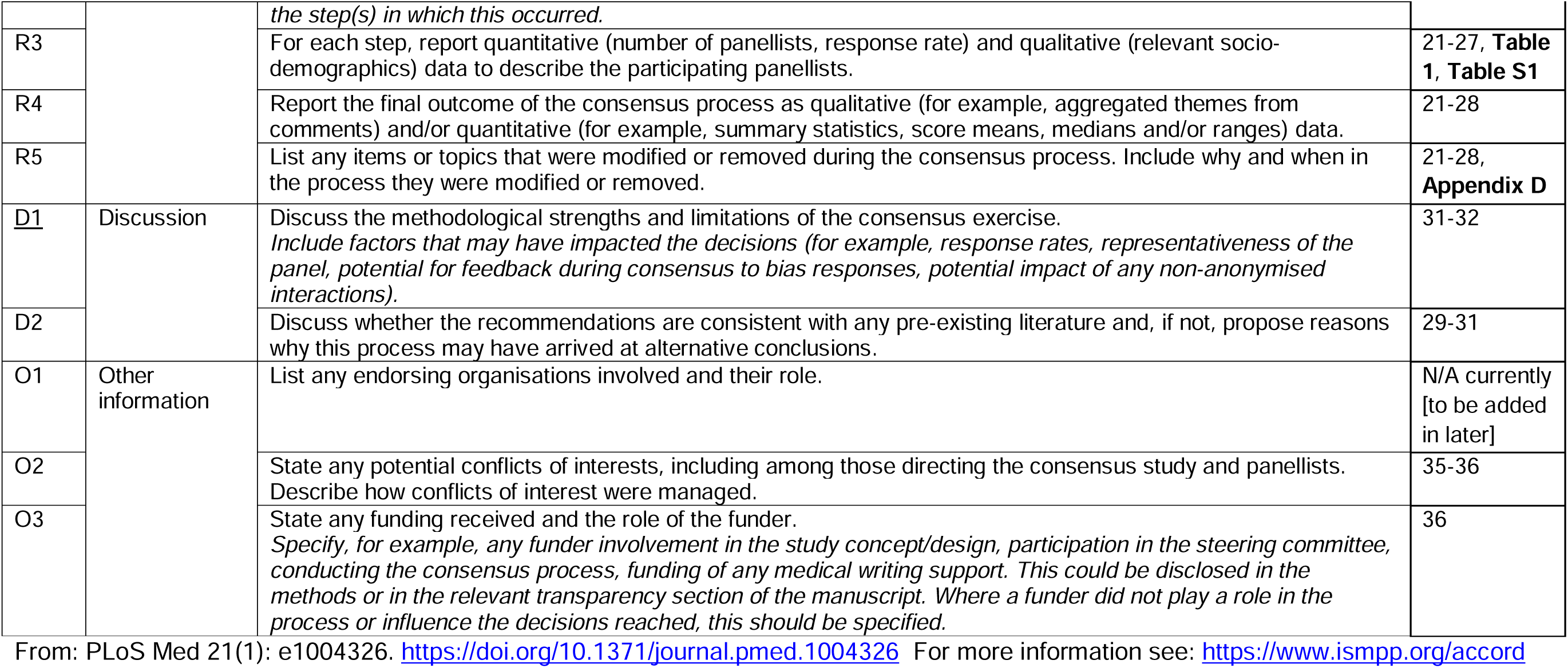
Accurate Consensus Reporting Document (ACCORD) Table.

**Table S2.**
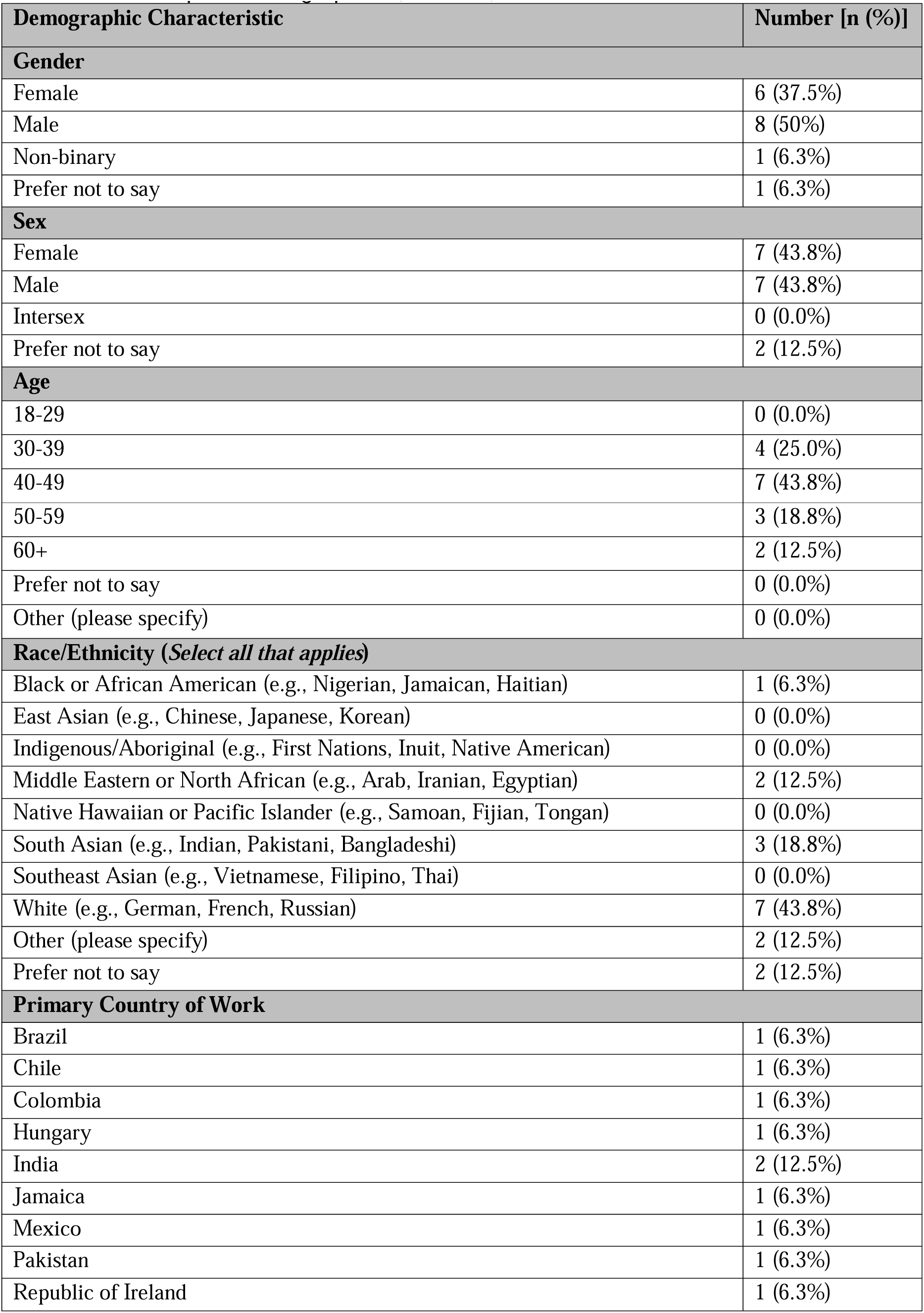
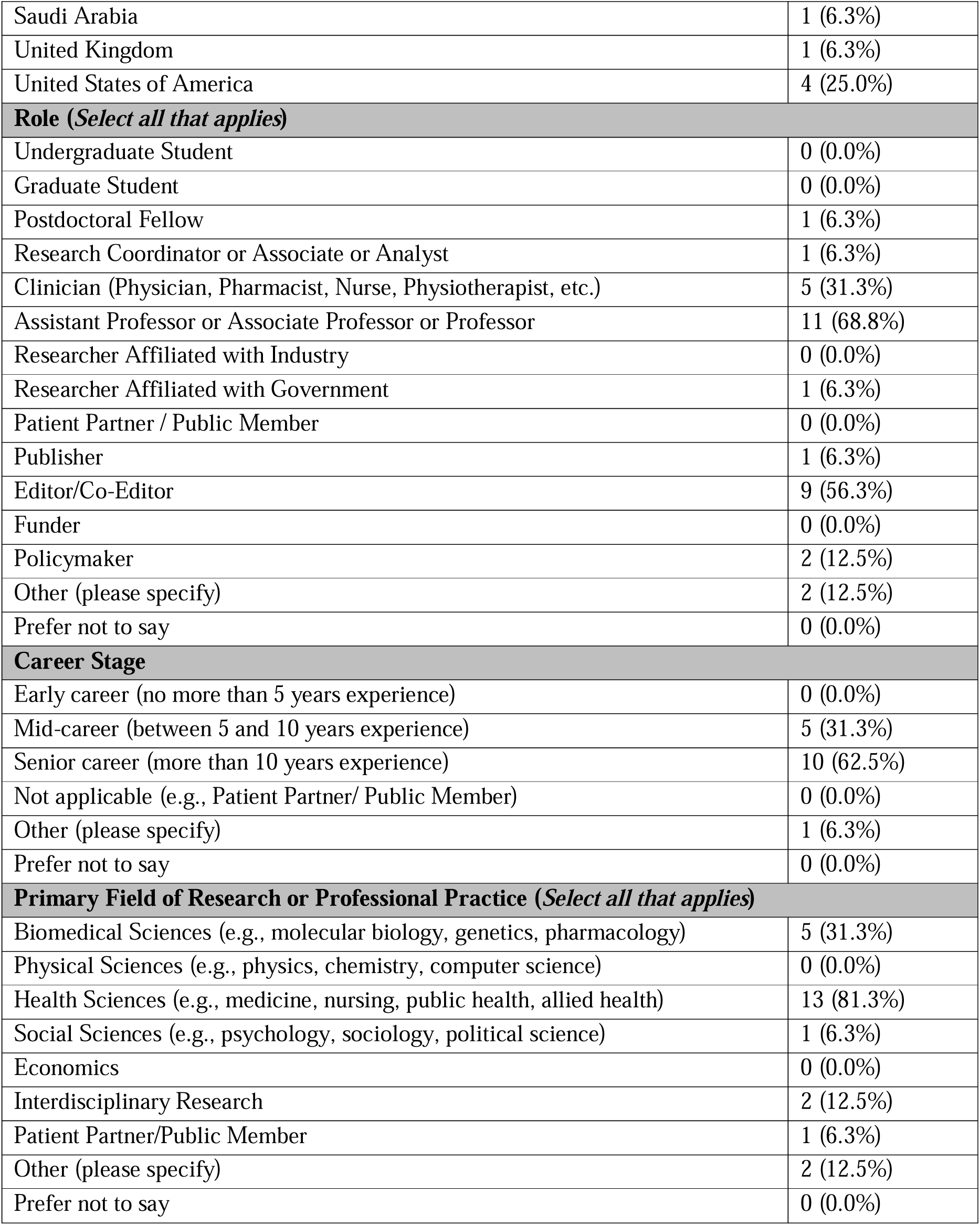
Participant Demographics (Round 3).

## References

1. Moxham N, Fyfe A. The Royal Society and the prehistory of peer review, 1665– 1965. Hist J 2018; 61: 863–889. DOI: 10.1017/S0018246X17000334.

2. Drozdz JA, Ladomery MR. The Peer Review Process: Past, Present, and Future. Br J Biomed Sci 2024; 81: 12054. DOI: 10.3389/bjbs.2024.12054.

3. Aczel B, Barwich A-S, Diekman AB, et al. The present and future of peer review: Ideas, interventions, and evidence. Proc Natl Acad Sci USA 2025; 122: e2401232121. DOI: 10.1073/pnas.2401232121.

4. Kelly J, Sadeghieh T, Adeli K. Peer Review in Scientific Publications: Benefits, Critiques, & A Survival Guide. EJIFCC 2014; 25: 227–243.

5. Bruce R, Chauvin A, Trinquart L, et al. Impact of interventions to improve the quality of peer review of biomedical journals: a systematic review and meta-analysis. BMC Med 2016; 14: 85. DOI: 10.1186/s12916-016-0631-5.

6. Grudniewicz A, Moher D, Cobey KD, et al. Predatory journals: no definition, no defence. Nature 2019; 576: 210–212. DOI: 10.1038/d41586-019-03759-y.

7. Wedderkopp N, Rutz E. Scientific Integrity and Transparency in Academic Writing: The Foundation of Credible Science. Children (Basel) 2024; 11: 1191. DOI: 10.3390/children11101191.

8. Zhaksylyk A, Zimba O, Yessirkepov M, et al. Research Integrity: Where We Are and Where We Are Heading. J Korean Med Sci 2023; 38: e405. DOI: 10.3346/jkms.2023.38.e405.

9. Alfonso F, Crea F. Preprints: a game changer in scientific publications? Eur Heart J 2023; 44: 171–173. DOI: 10.1093/eurheartj/ehac665.

10. Thibault RT, Amaral OB, Argolo F, et al. Open Science 2.0: Towards a truly collaborative research ecosystem. PLoS Biol 2023; 21: e3002362. DOI: 10.1371/journal.pbio.3002362.

11. Masood M, Tai KH, Rizzato JS, et al. Manuscript peer review lacks an empirically derived definition: a scoping review of 139 sources identifies 37 characteristics [In Press]. J Clin Epidemiol. DOI: 10.17605/OSF.IO/2E539.

12. Linstone HA, Turoff M. Delphi: A brief look backward and forward. Technological Forecasting and Social Change 2011; 78: 1712–1719. DOI: 10.1016/j.techfore.2010.09.011.

13. Dalkey N, Helmer O. An Experimental Application of the DELPHI Method to the Use of Experts. Management Science 1963; 9: 458–467. DOI: 10.1287/mnsc.9.3.458.

14. McMillan SS, King M, Tully MP. How to use the nominal group and Delphi techniques. Int J Clin Pharm 2016; 38: 655–662. DOI: 10.1007/s11096-016-0257-x.

15. Cobey KD, Lalu MM, Skidmore B, et al. What is a predatory journal? A scoping review. F1000Res 2018; 7: 1001. DOI: 10.12688/f1000research.15256.2.

16. Cukier S, Lalu M, Bryson GL, et al. Defining predatory journals and responding to the threat they pose: a modified Delphi consensus process. BMJ Open 2020; 10: e035561. DOI: 10.1136/bmjopen-2019-035561.

17. Wellcome Open Research, https://wellcomeopenresearch.org/.

18. Merton RK. The normative structure of science. In: The Sociology of Science: Theoretical and Empirical Investigations. Chicago, 1973, pp. 267–278.

19. OSF [Open Science Framework], https://osf.io/.

20. Masood M, Tai KH, Rizzato JS, et al. Defining Manuscript Peer Review: A Modified Delphi Consensus Process (Protocol), https://osf.io/z597r/files/ft2zx (2026).

21. medRxiv, https://www.medrxiv.org/.

22. Gattrell WT, Logullo P, Zuuren EJ van, et al. ACCORD (ACcurate COnsensus Reporting Document): A reporting guideline for consensus methods in biomedicine developed via a modified Delphi. PLOS Medicine 2024; 21: e1004326. DOI: 10.1371/journal.pmed.1004326.

23. Nakagawa S, Ivimey-Cook ER, Grainger MJ, et al. Method Reporting with Initials for Transparency (MeRIT) promotes more granularity and accountability for author contributions. Nat Commun 2023; 14: 1788. DOI: 10.1038/s41467-023-37039-1.

24. Akl EA, Khabsa J, Petkovic J, et al. “Interest-holders”: A new term to replace “stakeholders” in the context of health research and policy. Cochrane Evidence Synthesis and Methods 2024; 2: e70007. DOI: 10.1002/cesm.70007.

25. Cochrance Methods Equity. PROGRESS-Plus, https://methods.cochrane.org/equity/projects/evidence-equity/progress-plus.

26. Karran EL, Cashin AG, Barker T, et al. Using PROGRESS-plus to identify current approaches to the collection and reporting of equity-relevant data: a scoping review. J Clin Epidemiol 2023; 163: 70–78. DOI: 10.1016/j.jclinepi.2023.09.017.

27. Welphi, https://www.welphi.com/.

28. Abulela MAA, Khalaf MA. Does the Number of Response Categories Impact Validity Evidence in Self-Report Measures? A Scoping Review. Sage Open 2024; 14: 21582440241230363. DOI: 10.1177/21582440241230363.

29. Schifano J, Niederberger M. How Delphi studies in the health sciences find consensus: a scoping review. Syst Rev 2025; 14: 14. DOI: 10.1186/s13643-024-02738-3.

30. Zoom, https://www.zoom.com.

31. French SD, Green SE, O’Connor DA, et al. Developing theory-informed behaviour change interventions to implement evidence into practice: a systematic approach using the Theoretical Domains Framework. Implement Sci 2012; 7: 38. DOI: 10.1186/1748-5908-7-38.

32. Hsieh H-F, Shannon SE. Three Approaches to Qualitative Content Analysis. 15: 1277–1288. DOI: 10.1177/1049732305276687.

33. OpenAI. ChatGPT [Large language model], https://chatgpt.com/ (2026).

34. Thygesen K, Alpert JS, Jaffe AS, et al. Fourth Universal Definition of Myocardial Infarction (2018). J Am Coll Cardiol 2018; 72: 2231–2264. DOI: 10.1016/j.jacc.2018.08.1038.

35. Pinfield S, Waltman L, Kolarz P, et al. The Future of Peer Review: A system-wide perspective. Report, Research on Research Institute. DOI: 10.6084/m9.figshare.32685351.v1.

36. Reinhart M, Schendzielorz C. Peer-review procedures as practice, decision, and governance—the road to theories of peer review. Sci Public Policy 2024; 51: 543–552. DOI: 10.1093/scipol/scad089.

37. Silveira LD, Melero R, Caregnato SE, et al. Actitudes de los editores de revistas académicas brasileñas hacia la revisión por pares abierta (open peer review). Una encuesta. EPI 2023; e320620. DOI: 10.3145/epi.2023.nov.20.

38. Superchi C, Hren D, Blanco D, et al. Development of ARCADIA: a tool for assessing the quality of peer-review reports in biomedical research. BMJ Open 2020; 10: e035604. DOI: 10.1136/bmjopen-2019-035604.

39. Sever R. Biomedical publishing: Past historic, present continuous, future conditional. PLoS Biol 2023; 21: e3002234. DOI: 10.1371/journal.pbio.3002234.

40. COMET Intiative. Primary Evaluation outcomEs for Research on peer review – A Core Outcome Set (PEER-COS) [Registration], https://www.comet-initiative.org/Studies/Details/3934.

41. Masood M, Tai KH, Rizzato JS, et al. Appendix A_Round 1 Survey_9Aug2026, https://osf.io/w5gy8/files/euzdk (2026).

42. Masood M, Tai KH, Rizzato JS, et al. Appendix B_Round 2 Survey_9Aug2026, https://osf.io/w5gy8/files/hk936 (2026).

43. Masood M, Tai KH, Rizzato JS, et al. Appendix C_Round 3 Booklet_9Aug2026, https://osf.io/w5gy8/files/he927 (2026).

44. Masood M, Tai KH, Rizzato JS, et al. Appendix D_Quantitative Voting Item Results (All Rounds)_14Aug2026, https://osf.io/w5gy8/files/kj2nv (2026).

45. Masood M, Tai KH, Rizzato JS, et al. Appendix E_Qualitative Analysis Round 1_10Aug2026, https://osf.io/w5gy8/files/cuj48 (2026).

46. Masood M, Tai KH, Rizzato JS, et al. Appendix F_Qualitative Analysis Round 2_10Aug2026, https://osf.io/w5gy8/files/9pb7y (2026).

47. Masood M, Tai KH, Rizzato JS, et al. Appendix G_Round 1 Results (Complete Summarized)_9Aug2026, https://osf.io/w5gy8/files/a8xuk (2026).

48. Masood M, Tai KH, Rizzato JS, et al. Appendix H_Round 2 Results (Complete Summarized)_9Aug2026, https://osf.io/w5gy8/files/6xb3f (2026).

49. Masood M, Tai KH, Rizzato JS, et al. Appendix I_Round 3 Results (Complete Summarized)_14Aug2026, https://osf.io/w5gy8/files/ec24m (2026).

