## Supplementary figures and images for "Defining Manuscript Peer Review: An International Delphi Consensus Study"

### Manuscript

**Figure 1.** Participants Flowchart.
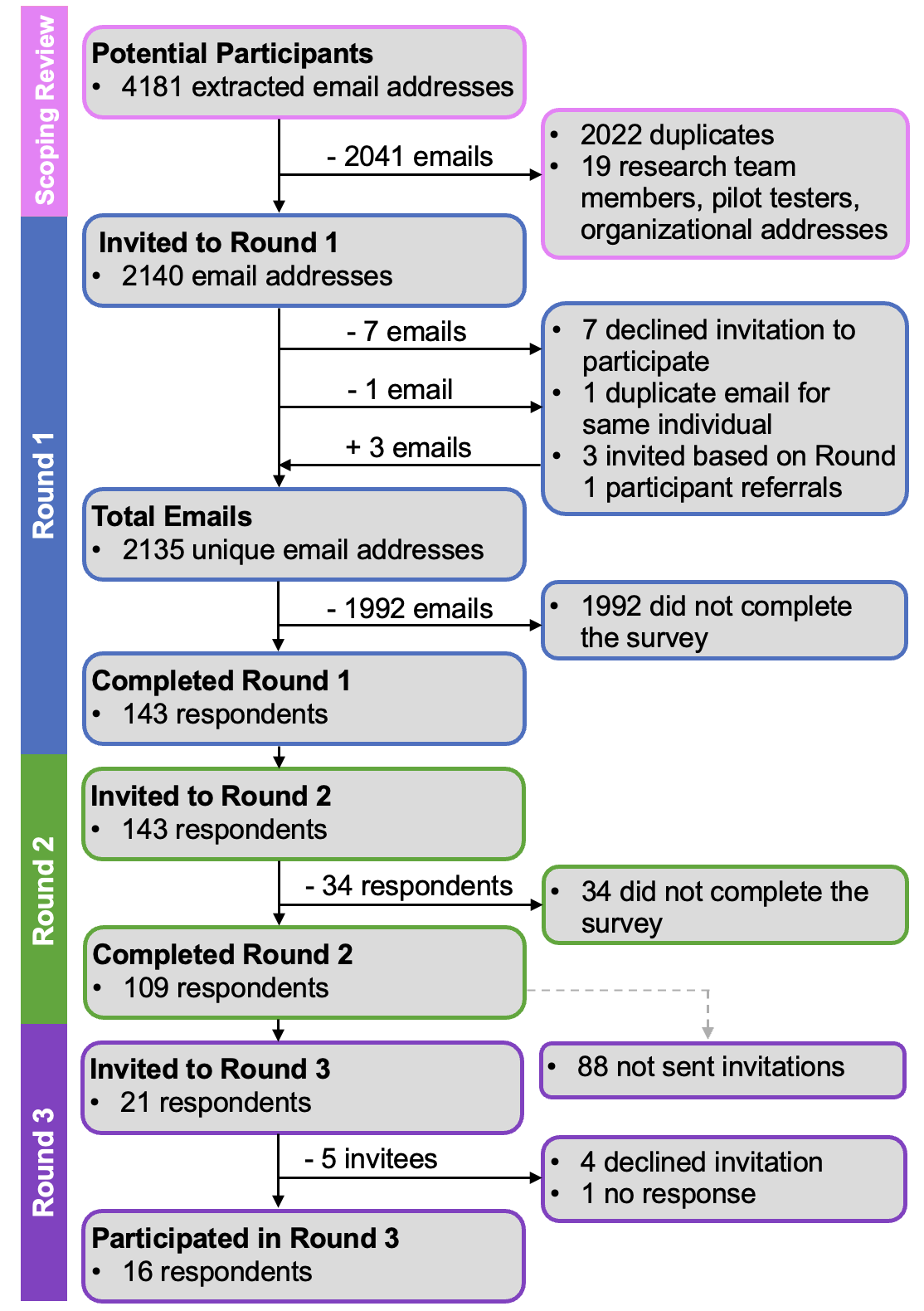

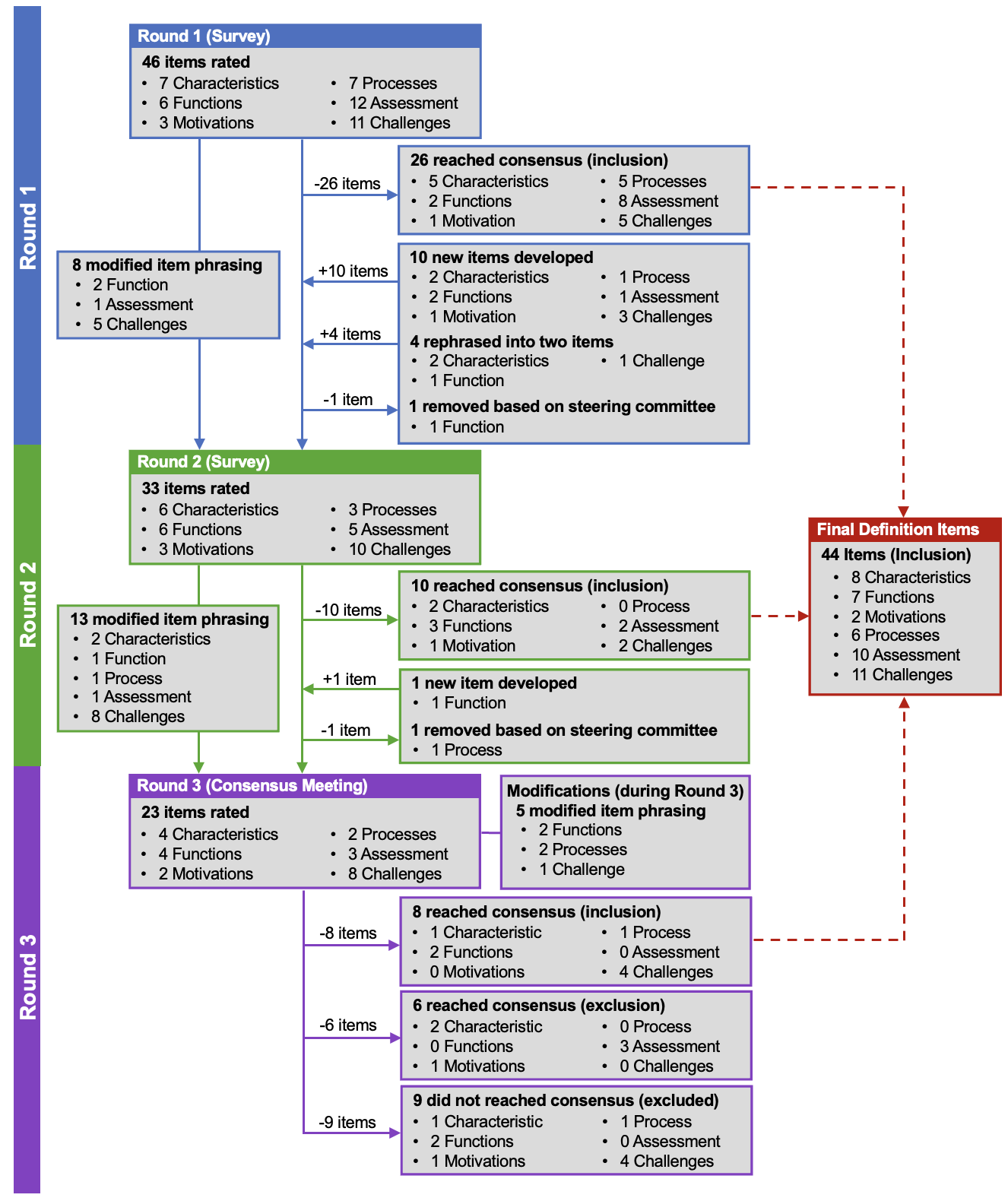
**Figure 2.** Peer Review Features: Item Tracking Flowchart.
