## Supplementary material for "Defining Manuscript Peer Review: An International Delphi Consensus Study": Figures

### Tables

**Table 1.** Participant Demographics (Rounds 1 and 2).

| **Demographic Characteristics** | **Number [n (%)]** | |
| --- | --- | --- |
|  | **Round 1  (N = 143)** | **Round 2**  **(N = 109)** |
| **Gender** | | |
| Female | 48 (33.6%) | 33 (30.3%) |
| Male | 92 (64.3%) | 73 (67.0%) |
| Non-binary | 1 (0.7%) | 1 (0.9%) |
| Prefer not to say | 2 (1.4%) | 2 (1.8%) |
| **Sex at birth** | | |
| Female | *Not asked in Round 1* | 34 (31.2%) |
| Male |  | 72 (66.1%) |
| Intersex |  | 0 (0.0%) |
| Prefer not to say |  | 3 (2.8%) |
| **Age** | | |
| 18-29 | 2 (1.4%) | 1 (0.9%) |
| 30-39 | 20 (14.0%) | 18 (16.5%) |
| 40-49 | 38 (26.6%) | 29 (26.6%) |
| 50-59 | 30 (21.0%) | 21 (19.3%) |
| 60+ | 52 (36.4%) | 39 (35.8%) |
| Prefer not to say | 1 (0.7%) | 1 (0.9%) |
| Other (please specify) | 4 (2.8%) | 1 (0.9%) |
| **Race/Ethnicity (*Select all that applies*)** | | |
| Black or African American (e.g., Nigerian, Jamaican, Haitian) | 1 (0.7%) | 1 (0.9%) |
| East Asian (e.g., Chinese, Japanese, Korean) | 6 (4.2%) | 3 (2.8%) |
| Indigenous/Aboriginal (e.g., First Nations, Inuit, Native American) | 0 (0.0%) | 0 (0.0%) |
| Middle Eastern or North African (e.g., Arab, Iranian, Egyptian) | 4 (2.8%) | 4 (3.7%) |
| Native Hawaiian or Pacific Islander (e.g., Samoan, Fijian, Tongan) | 1 (0.7%) | 1 (0.9%) |
| South Asian (e.g., Indian, Pakistani, Bangladeshi) | 13 (9.1%) | 12 (11.0%) |
| Southeast Asian (e.g., Vietnamese, Filipino, Thai) | 1 (0.7%) | 1 (0.9%) |
| White (e.g., German, French, Russian) | 109 (76.2%) | 81 (74.3%) |
| Other (please specify) | 8 (5.6%) | 7 (6.4%) |
| Prefer not to say | 7 (4.9%) | 6 (5.5%) |
| **Primary Country of Work** | | |
| United States of America | 50 (35.0%) | 43 (39.4%) |
| United Kingdom | 19 (13.3%) | 10 (9.2%) |
| Canada | 7 (4.9%) | 3 (2.8%) |
| India | 7 (4.9%) | 7 (6.4%) |
| Australia | 6 (4.2%) | 6 (5.5%) |
| Germany | 6 (4.2%) | 4 (3.7%) |
| Italy | 6 (4.2%) | 4 (3.7%) |
| Other | 42 (29.4%) | 66 (60.6%) |
| **Role (*Select all that applies*)** | | |
| Undergraduate Student | 0 (0.0%) | 0 (0.0%) |
| Graduate Student | 1 (0.7%) | 1 (0.9%) |
| Postdoctoral Fellow | 6 (4.2%) | 5 (4.6%) |
| Research Coordinator or Associate or Analyst | 7 (4.9%) | 5 (4.6%) |
| Clinician (Physician, Pharmacist, Nurse, Physiotherapist, etc.) | 36 (25.2%) | 29 (26.6%) |
| Assistant Professor or Associate Professor or Professor | 81 (56.6%) | 63 (57.8%) |
| Researcher Affiliated with Industry | 4 (2.8%) | 1 (0.9%) |
| Researcher Affiliated with Government | 8 (5.6%) | 8 (7.3%) |
| Patient Partner / Public Member | 0 (0.0%) | 0 (0.0%) |
| Publisher | 8 (5.6%) | 6 (5.5%) |
| Editor/Co-Editor | 55 (38.5%) | 43 (39.4%) |
| Funder | 1 (0.7%) | 1 (0.9%) |
| Policymaker | 2 (1.4%) | 2 (1.8%) |
| Other (please specify) | 23 (16.1%) | 17 (15.6%) |
| Prefer not to say | 1 (0.7%) | 1 (0.9%) |
| **Career Stage** | | |
| Early career (no more than 5 years experience) | 9 (6.3%) | 7 (6.4%) |
| Mid-career (between 5 and 10 years experience) | 18 (12.6%) | 17 (15.6%) |
| Senior career (more than 10 years experience) | 111 (77.6%) | 80 (73.4%) |
| Not applicable (e.g., Patient Partner/ Public Member) | 1 (0.7%) | 1 (0.9%) |
| Other (please specify) | 4 (2.8%) | 4 (3.7%) |
| Prefer not to say | 0 (0.0%) | 0 (0.0%) |
| **Primary Field of Research or Professional Practice (*Select all that applies*)** | | |
| Biomedical Sciences (e.g., molecular biology, genetics, pharmacology) | 35 (24.5%) | 24 (22.0%) |
| Physical Sciences (e.g., physics, chemistry, computer science) | 2 (1.4%) | 2 (1.8%) |
| Health Sciences (e.g., medicine, nursing, public health, allied health) | 91 (63.6%) | 73 (67.0%) |
| Social Sciences (e.g., psychology, sociology, political science) | 22 (15.4%) | 15 (13.8%) |
| Economics | 2 (1.4%) | 1 (0.9%) |
| Interdisciplinary Research | 16 (11.2%) | 12 (11.0%) |
| Patient Partner/Public Member | 2 (1.4%) | 2 (1.8%) |
| Other (please specify) | 12 (8.4%) | 11 (10.1%) |
| Prefer not to say | 0 (0.0%) | 0 (0.0%) |
| **Total Manuscripts Submitted for Publication (as an author)** | | |
| 0 | 0 (0.0%) | 0 (0.0%) |
| <10 | 10 (7.0%) | 6 (5.5%) |
| 10–19 | 11 (7.7%) | 8 (7.3%) |
| 20–29 | 6 (4.2%) | 5 (4.6%) |
| 30–39 | 9 (6.3%) | 7 (6.4%) |
| 40–49 | 2 (1.4%) | 2 (1.8%) |
| >50 | 103 (72.0%) | 79 (72.5%) |
| Other (please specify) | 2 (1.4%) | 2 (1.8%) |
| **Experience Reviewing Manuscripts (as a peer reviewer)** | | |
| None | 1 (0.7%) | 1 (0.9%) |
| <1 year | 4 (2.8%) | 1 (0.9%) |
| 1–5 years | 8 (5.6%) | 7 (6.4%) |
| 6–10 years | 19 (13.3%) | 15 (13.8%) |
| >10 years | 109 (76.2%) | 83 (76.1%) |
| Unsure | 1 (0.7%) | 1 (0.9%) |
| Other (please specify) | 1 (0.7%) | 1 (0.9%) |
| **Number of Manuscripts Reviewed in last 5 years (as a peer reviewer)** | | |
| None | 2 (1.4%) | 2 (1.8%) |
| <10 | 15 (10.5%) | 10 (9.2%) |
| 10–20 | 17 (11.9%) | 12 (11.0%) |
| 21–50 | 38 (26.6%) | 29 (26.6%) |
| >50 | 66 (46.2%) | 51 (46.8%) |
| Unsure | 1 (0.7%) | 1 (0.9%) |
| Other (please specify) | 4 (2.8%) | 4 (3.7%) |
| **Formal Training in Manuscript Peer Review** | | |
| Yes | 69 (48.3%) | 59 (54.1%) |
| No | 69 (48.3%) | 47 (43.1%) |
| Don’t remember | 3 (2.1%) | 2 (1.8%) |
| Other (please specify) | 2 (1.4%) | 1 (0.9%) |
| **Type of Manuscript Peer Review Training (*Select all that applies*)** | | |
| University/Graduate course | 7 (4.9%) | 5 (4.6%) |
| Journal- or publisher-based training program | 36 (25.2%) | 31 (28.4%) |
| Online course or webinar | 17 (11.9%) | 15 (13.8%) |
| In-person workshop or seminar | 22 (15.4%) | 19 (17.4%) |
| Mentorship/Apprenticeship from a senior researcher | 27 (18.9%) | 25 (22.9%) |
| Self-directed training (e.g., reading guidelines, online resources) | 59 (41.3%) | 48 (44.0%) |
| Other (please specify) | 4 (2.8%) | 3 (2.8%) |
| Not applicable (I did not receive training) | 69 (48.3%) | 48 (44.0%) |

^a^ A complete summary of all answers, including for the ‘Other (please specify)’ sections, are provided in **Appendix G** and **Appendix H** for Rounds 1 and 2 respectively.

**Table 2.** Consensus Items (inclusion).

| **Item** | **Consensus Round*** | **Participants**  **[n (%)†]** |
| --- | --- | --- |
| **Characteristics** | | |
| **C1. Principled:** Peer review should be principled  (E.g., Honest, fair, objective, transparent, trustworthy, moral, incorruptible, open-minded, unbiased) | 1 | 139 (97.2%) |
| **C2. Constructive:** Peer review should be constructive (E.g., Aiming at improving manuscripts, corrective, actionable) | 1 | 134 (93.7%) |
| **C3. Timely:** Peer review should be given an expected date to be completed by | 1 | 126 (88.1%) |
| **C4. Professional:** Peer review should be professional (E.g., Respectful, supportive, interactive, diplomatic, kind, dialogical, community-building) | 1 | 138 (96.5%) |
| **C5. Quality:** Peer review should be critical (E.g., Competent, evidence-based, well-reasoned, concrete, well-organised, thorough, knowledgeable) | 1 | 140 (97.9%) |
| **C6. Confidential:** Manuscript under review should be conditionally confidential (E.g., Protecting the yet-to-be-published research from early disclosure) | 2 | 92 (84.4%) |
| **C7. Transparent:** Peer review process and content should be transparent (E.g., Peer reviewer selection, peer review timeline, peer review decision) | 2 | 94 (86.2%) |
| **C8. Pluralistic:** When feasible, peer review should be conducted with multiple reviewers to incorporate diverse perspectives  (E.g., gathering opinions from different perspectives and reviewers with geographical and demographical diversity) | 3 | 13 (81.3%) |
| **Functions** | | |
| F1. Peer review safeguards publication by improving the quality of accepted manuscripts | 1 | 123 (86.0%) |
| F2. Peer review improves science by strengthening the trustworthiness and reliability of knowledge | 1 | 128 (89.5%) |
| F3. Peer review is intended to safeguard publication by filtering out suboptimal manuscripts | 2 | 99 (90.8%) |
| F4. Peer review is intended to improve science by safeguarding research ethics and integrity | 2 | 101 (92.7%) |
| F5. Peer review is intended to provide feedback to improve the completeness and transparency of author's manuscripts (e.g., clear justification of accept/reject decisions, actionable feedback for authors) | 2 | 102 (93.6%) |
| F6. Peer review is intended to improve the academic credibility of manuscripts | 3 | 15 (93.8%) |
| F7. Peer review is intended to facilitate scholarly discussion | 3 | 16 (100.0%) |
| **Motivations** | | |
| M1. Peer review participation is motivated by a sense of duty to the community (E.g., Communal norms of duty, reciprocity, service to the scholarly community, collective responsibility) | 1 | 127 (88.8%) |
| M2. Peer review participation is motivated by community gains (E.g., Research quality betterment, information exchange, community building) | 2 | 95 (87.2%) |
| **Process** | | |
| P1. Peer review involves editors selecting peer reviewers and matching their expertise (E.g., Subject-matter expertise, current disciplinary knowledge, methodological competence) | 1 | 131 (91.6%) |
| P2. Peer review involves reviewers providing evaluative comments on manuscript (E.g., Strengths, weaknesses, suggestions, revision requirements) | 1 | 143 (100.0%) |
| P3. Peer review involves editors aggregating and weighing multiple independent reviews | 1 | 129 (90.2%) |
| P4. Peer review involves editors deciding on the acceptance of a manuscript | 1 | 122 (85.3%) |
| P5. Peer review proceeds through iterative rounds of review and revision | 1 | 114 (79.7%) |
| P6. Peer review involves reviewers advising on the suitability for publication to editors | 3 | 12 (80.0%) |
| **Assessment** | | |
| A1. Peer review evaluates the overall quality of manuscripts (E.g., Overall clarity, structure, completeness, logical flow, consistency, language) | 1 | 138 (96.5%) |
| A2. Peer review evaluates the quality of research (E.g., Scientific rigor, research quality, finding validity) | 1 | 139 (97.2%) |
| A3. Peer review evaluates the originality of research (E.g., Research novelty, contribution of research to the existing literature) | 1 | 124 (86.7%) |
| A4. Peer review evaluates the relevance of research to its field (E.g., Importance of research, contribution to the community, trending topic, gaps in the field) | 1 | 132 (92.3%) |
| A5. Peer review evaluates study design and methods (E.g., Research question, sampling, data collection, data source, confounding controls) | 1 | 140 (97.9%) |
| A6. Peer review evaluates reporting quality (E.g., Adherence to reporting guidelines, clarity, internal consistency, transparency, interpretive appropriateness) | 1 | 133 (93.0%) |
| A7. Peer review evaluates statistical analysis (E.g., Analytic method, modelling, fitness of model, sample size) | 1 | 128 (89.5%) |
| A8. Peer review evaluates literature cited within the manuscript (E.g., Coverage, currency, referencing quality, citation bias) | 1 | 126 (88.1%) |
| A9. Peer review evaluates the transparency of reporting research (E.g., Transparency and reproducibility, research material availability, data and code sharing, result dissemination, protocol registration, trial pre-registration) | 2 | 94 (86.2%) |
| A10. Peer review evaluates the ethics and integrity of research (E.g., Declaration of conflict of interests, funding transparency, acknowledgements, ethical approval, participant consent, questionable research practices) | 2 | 102 (93.6%) |
| **Challenges** | | |
| Ch1. The process of peer review is time-consuming (E.g., Prolonged and unpredictable timelines, iterative review cycles) | 1 | 123 (86.0%) |
| Ch2. Peer review is limited by low availability of reviewers (E.g., Reviewer scarcity, low response rates of reviewers, lack of topic-specific expert in niche field) | 1 | 129 (90.2%) |
| Ch3. Peer review is limited by low incentive of reviewers (E.g., Unpaid, voluntary, and under-recognised labour of reviewers) | 1 | 118 (82.5%) |
| Ch4. The workload of peer review is demanding (E.g., Overloading reviewers, uneven workload distribution) | 1 | 127 (88.8%) |
| Ch5. Peer review is prone to biases (E.g., Author, reviewer, editor, journal, and field bias; Related to gender, geography, ethnics, status, language, ideology, career stage) | 1 | 116 (81.1%) |
| Ch6. Peer review can be affected by ethical and quality pitfalls (E.g., Coercive citation, ghostwriting of review, conflict of interest, substandard review, unsolicited comments, inappropriate delay in publication, biases of reviewers) | 2 | 104 (95.4%) |
| Ch7. Peer review is challenged by uncertainty regarding the appropriate use of AI | 2 | 90 (82.6%) |
| Ch8. Quality assurance in peer review is inconsistent across journals | 3 | 12 (80.0%) |
| Ch9. Power imbalances between authors, reviewers, and editors can influence peer review (E.g., seniority, reputation, institutional prestige, editorial authority) | 3 | 14 (93.3%) |
| Ch10. Academic reward systems do not adequately recognise reviewer contributions | 3 | 13 (86.7%) |
| Ch11. Securing reviewers with appropriate expertise and competence is challenging | 3 | 13 (86.7%) |
| **Reviewer Characteristics** | | |
| R1. Same or related discipline | 1 | 123 (86.0%) |
| R2. A peer must be a human  (i.e., it cannot be a computer/AI algorithm) | 1 | 116 (81.1%) |
| R3. Have shared expertise in the same methodology as the authors | 2 | 89 (81.7%) |
| R4. Have similar interests or experience in the research topic of the manuscript | 2 | 90 (82.6%) |
| R5. Be willing and able to dedicate sufficient time to complete manuscript peer review | 3 | 15 (100.0%) |
| R6. Be objective when evaluating manuscripts | 3 | 12 (80.0%) |

* Defines the round in which consensus was achieved. The three Delphi rounds included 143 participants in Round 1, 109 in Round 2, and 16 in Round 3.

† The reported n (%) values represent the number and proportion of participants meeting the criterion for inclusion in each consensus round. In Rounds 1 and 2, consensus for inclusion was defined as ≥80% (rounded to the nearest integer) of respondents rating an item within the upper third of the 5-point scale (scores 4–5). In Round 3, consensus for inclusion was defined as ≥80% of participants selecting “include” among those selecting either “include” or “exclude”; participants selecting “abstain” were excluded from the denominator.
